# Population health and health sector cost impacts of the UK Soft Drinks Industry Levy: a modelling study

**DOI:** 10.1101/2023.10.05.23296619

**Authors:** Linda J Cobiac, Cherry Law, Richard Smith, Steven Cummins, Harry Rutter, Mike Rayner, Oliver Mytton, Adam D M Briggs, Henning Tarp Jensen, Marcus Keogh-Brown, Jean Adams, Martin White, Peter Scarborough

**Author notes:** Correspondence to: Peter Scarboourough.

## Abstract

**Objective:** To model future impacts of the UK Soft Drinks Industry Levy (SDIL) on population health and health sector costs, and to estimate net monetary benefit to the health system.

**Design:** Proportional multi-state lifetable modelling study

**Setting:** United Kingdom

**Population:** All children and adults

**Intervention:** The SDIL is a two-tier levy of £0.18 per litre on drinks with between 5g and 8g of total sugars per 100mL, and £0.24 per litre on drinks with 8g or more of total sugars per 100mL. We estimated a per person reduction in sugar from previous interrupted time series analysis, which found an 8.0 g/household/week (95% confidence interval: 2.4 to 13.6) reduction in sugar in purchased drinks at one year after implementation.

**Main outcome measures:** *W*e evaluated impact of the sugar reduction on: (a) prevalence of overweight and obesity, obesity-related diseases and dental health out to 2050; and (b) lifetime population health (measured in quality-adjusted life years [QALYs]), change in costs to the health sector and the resulting net monetary benefit.

**Results:** The model predicts that the SDIL will reduce prevalence of overweight and obesity in the UK by 0.18 percentage points (95% uncertainty interval: 0.059 to 0.31) for males and 0.20 percentage points (0.064 to 0.34) for females, for as long as the sugar-reduction effects of the SDIL are sustained. In the first ten years of implementation, the reductions in sugar and overweight/obesity are predicted to prevent 270,000 (35,000 to 600,000) dental caries, 12,000 (3,700 to 20,000) cases of type 2diabetes, 3,800 (1,200 to 6,700) cases of cardiovascular diseases, and 350 (110 to 590) cases of obesity-related cancer. For the current UK population, it is estimated the SDIL will add 200,000 QALYs (63,500 to 342,000) over their lifetime and avert £174 million (£53.6 to £319) in their costs of health care (discounted at UK Treasury rates). At a UK Treasury value of £60,000 per QALY, it is estimated the SDIL will produce a net monetary benefit of £12.2 billion (£3.88 to £20.8) for the health system.

**Conclusion:** This study of the UK SDIL tiered tax on sugar content provides further evidence that sugar- sweetened beverage taxes have the potential to achieve meaningful improvements in population health and reduce health sector spending.

**What is already known on this topic:**

- Numerous modelling studies have shown that sugar-sweetened beverage taxes are likely to be cost-effective and improve population health, whether the tax is applied on the volume of product or to the sugar content (absolute or tiered), but there have been few modelling studies that have been informed by evaluation of real-world taxes.
- A volumetric sugar-sweetened beverage tax of 1 peso per litre, implemented in Mexico in 2014, reduced purchases of sugar-sweetened beverages by 7.6% in the first two years, and health economic modelling estimated that it would be cost-effective from a health sector perspective.
- In the UK, the Soft Drinks Industry Levy (SDIL), announced in March 2016 and implemented in April 2018, was designed as a tiered levy to encourage soft drink manufacturers to reduce sugar content. Follow-up at one year indicates that it has reduced purchases of sugar from drinks by 8.0 g/household/week (95% confidence interval: 2.4 to 13.6).

**What this study adds:**

- Population health modelling suggests that changes in sugar consumption due to the SDIL will reduce prevalence of overweight/obesity and related diseases and improve dental health in the UK, including 12,000 (95% uncertainty interval: 3,700 to 20,000) fewer cases of type 2 diabetes, 3,800 (1,200 to 6,700) fewer cases of cardiovascular diseases, 350 (110 to 590) fewer cases of obesity-related cancer, and 270,000 (35,000 to 600,000) fewer dental caries, in the first ten years after implementation.
- Health economic modelling indicates that over the lifetime of the current UK population the SDIL could add 200,000 quality-adjusted life years (63,500 to 342,000) and avert £174 million (£53.6 to £319) in health care costs, leading to a net monetary benefit of £12.2 billion (£3.88 to £20.8) for the health sector.
- This study of the UK SDIL tiered tax on sugar content provides further evidence that sugar- sweetened beverage taxes have the potential to achieve meaningful improvements in population health and reduce health sector spending.

## Introduction

In 2016 the UK Chancellor of the Exchequer announced that the UK Government would implement incentives for soft drink manufacturers, importers and bottlers to reduce the amount of sugar in soft drinks in the UK.^1^ The Soft Drinks Industry Levy (SDIL) was implemented in 2018 as a two tier levy of £0.18 per litre on drinks with between 5g and 8g of total sugars per 100mL, and £0.24 per litre on drinks with 8g or more of total sugars per 100mL.^2^ The levy targets soft drinks where sugar or similar sweeteners, such as honey, are added during the manufacturing process. Fruit or vegetable juices with no added sugar, and drinks that are at least 75% milk, among others, are excluded.

Sugar-sweetened beverages are a rational target for a levy, since they provide no nutritional benefit in the diet. Experimental studies have shown that they are associated with weight gain in children.^3^ ^4^ Those who regularly consume these drinks are more likely to have overweight or obesity, and experience type 2 diabetes and cardiovascular disease.^5–9^ The low impact of these drinks on feelings of satiety, and subsequent over-consumption of calories, is understood to be a key mechanism underlying these effects, although these is also emerging evidence of other adverse metabolic mechanisms.^10^ Oral health is also affected, with greater consumption of sugar-sweetened beverages associated with increased risk of dental caries.^11^

Previous modelling analyses predicted that a sugar-sweetened beverage tax in the UK would improve population health by reducing prevalence of obesity and rates of cardiovascular disease, diabetes, cancer and dental caries.^12^ ^13^ But these studies relied on speculation of the likely effects on product reformulation and changes in market share of products with different levels of sugar content, and relied on price elasticities to estimate consumer responses to potential changes in price. Following implementation of the SDIL in the UK, we now have the opportunity to examine the population health and health care cost implications of the tax based on real-world evaluation of the effects on sugar content and drink purchasing. While many countries have now implemented sugar-sweetened beverage taxes,^14^ to date only in Mexico has evidence from a real-world evaluation been used to examine the future consequences for population health and health care expenditure. In the first two years after implementation of a 1 peso per litre tax in Mexico in 2014, purchases of sugar-sweetened beverages reduced by an average of 7.6%.^15^. Health economic modelling estimated that the tax would be cost-effective from a health sector perspective, predicting significant reductions in prevalence of obesity and cases of diabetes, cardiovascular disease and cancers, and reduced health care expenditure.

In this study, we model the health implications of the SDIL in the UK, based on real-world evaluation of its effects on the sugar content and purchasing of soft drinks. Interrupted time series analyses of purchasing trends before and after the announcement and implementation of the SDIL found that at one year after implementation the purchased volume of soft drinks had increased by 188.8 mL/household/week (95% confidence interval: 30.7 to 346.9) but that the amount of sugar in purchased drinks had decreased by 8.0 g/household/week (13.6 to 2.4 g).^16^ We model the future impacts of this sugar reduction on obesity, diabetes, cardiovascular disease, cancer and dental health in the UK population, and the cost implications of these changes for the health sector.

## Methods

We modelled the health and health care cost impacts of the SDIL using PRIMEtime,^17^ a proportional multi-state lifetable model developed in the UK for simulating population health and cost impacts of public health interventions and scenarios. PRIMEtime simulates the changing health of a population through time. From changes in prevalence of behavioural risk factors, such as obesity, the model can simulate impacts on incidence, prevalence and mortality of non-communicable diseases, including cardiovascular diseases, diabetes and cancers. These changes in disease influence both the years of life that are lived by the population and their quality of life, from which we can estimate the impact of an intervention or scenario on life expectancy, quality-adjusted life years (QALYs) and health sector costs (Figure 1). A full description of the PRIMEtime input data for these analyses are shown in Text S1.

**Figure 1.**
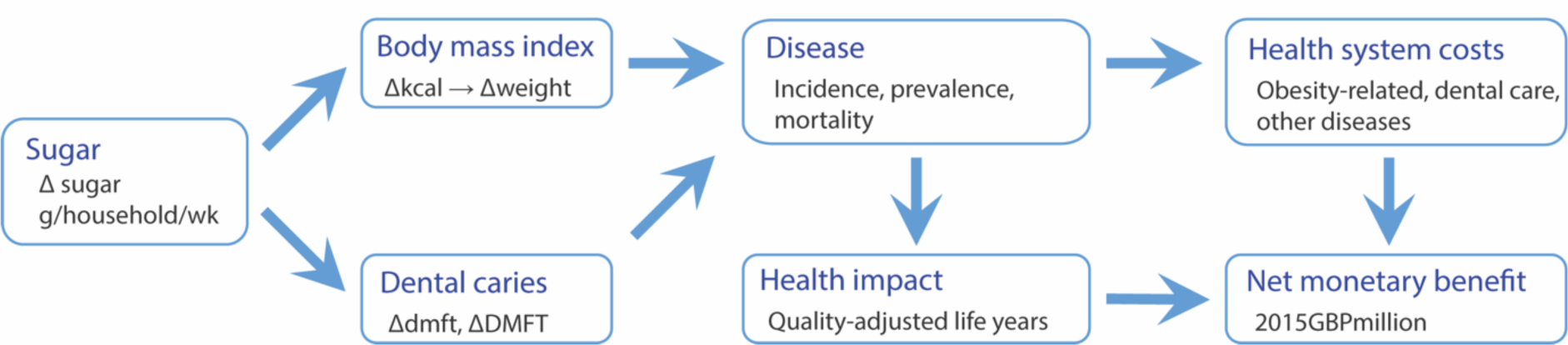
Modelling the effects of the SDIL on population health and health system costs (NB. dmft/DMFT – decayed missing and filled deciduous/permanent teeth)

### SDIL effect on sugar purchasing

We derived the effect of the SDIL on purchased sugar in drinks from the results of an interrupted time series analysis.^10^ This evaluation study analysed commercial household purchasing panel data from before announcement of the SDIL (March 2014-March 2016) to one year post-implementation (April 2018-March 2019). The data included purchases of drinks eligible for the levy (e.g. soft drinks) and drinks that were exempt (e.g. milk, milk-based drinks, alcoholic drinks, no-added-sugar fruit juices and drinks sold as powders), to capture potential substitution effects. The panel data only included purchases for consumption at home; drinks purchased and consumed outside the home (e.g. as part of a meal, on a journey, or at a restaurant) were excluded. The study found a net reduction in purchased sugar of 8.0 g/household/week (13.6 to 2.4 g).^16^ From this, we estimated an average per person reduction in sugar consumption, assuming an average of 2.4 people per household,^18^ and assuming that changes in purchases translate directly into changes in dietary sugar intake.^19^ ^20^ We did not assume any variation in modelled consumption by age or sex, since examination of National Diet and Nutrition Survey data in the years before the announcement of the SDIL did not indicate any observable variation by age or sex in the consumption of sugar from across the range of included drinks (soft drinks, bottled water, milk and milk-based drinks, no-added-sugar juices and drinks sold as powders).^21^

### Dental caries

We defined dental caries by the number of decayed, missing and filled permanent (DMFT) or deciduous (dmft) teeth. In the absence of UK-specific data, we estimated the effect of the sugar reduction on dental caries, based on the results of a longitudinal analysis of sugar consumption and dental caries in three national surveys (2000, 2004 and 2011) in Finland.^12^ ^22^ In this study, Bernabe et al found an increase of 0.09 DMFT (95% CI: 0.02 to 0.15) for each additional 10g of sugar consumed each day over the 11 year period, in the study population aged 30+ years. Given a substantial body of evidence demonstrating a relationship between the amount of sugar consumed and development of dental caries in both children and adults,^23^ we assumed the dose-response relationship applied equally to dmft.

For children, we determined current dmft and DMFT rates from the Children’s Dental Health Survey 2013.^24^ For adults, we derived DMFT rates from separately measured numbers of decayed, missing and filled teeth reported in the Adult Dental Health Survey 2009,^25^ adjusting for congenital tooth absence, tooth extraction from causes other than dental decay (e.g. periodontal disease, trauma) and edentulism.^26–28^

### Body mass index

We estimated the effect of sugar reduction on body mass index (BMI) using energy balance equations for children and adults,^29^ ^30^ assuming 3.75 kcal per gram of sugar,^31^ and no compensatory changes in energy expenditure or substitution of calories from other foods or drinks. Carbohydrates consumed in liquid form do not have the same effects on feelings of satiety that they do when consumed in solid food,^32^ ^33^ which may lessen the desire to increase consumption and calorie intake from other foods or drinks. Studies of sugar-sweetened beverage taxes do not indicate compensatory increases in food purchasing.^34^ ^35^ In the evaluation of the SDIL, there was evidence of increases in purchasing of low or no sugar drinks, due to reformulation (which are accounted for in our modelling), but there was no evidence of an increase in purchasing of confectionery (a potential substitute for sugar and/or calorie intake) over the time period of announcement and implementation of the SDIL.^36^

We modelled the SDIL effect on BMI as a change in trajectory of the shape and scale of the lognormal distribution of BMI, by age and sex, based on prediction models developed and validated on 27 years of Health Survey for England data.^37^ For the main analyses we used non-linear prediction models that forecast mean BMI heading towards an asymptote. In sensitivity analyses we used linear prediction models that forecast mean BMI reaching a peak within the next decade and declining thereafter. Prevalence of overweight and obesity were defined using BMI thresholds from the International Obesity Taskforce.^38^

### Disease modelling

We evaluated the impact of BMI changes on the incidence of five cardiovascular and metabolic diseases (ischaemic heart disease, ischaemic stroke, intracerebral haemorrhage, hypertensive heart disease atrial fibrillation/flutter, and diabetes mellitus type 2) and eight obesity-related cancers (colorectal cancer, post-menopausal breast cancer, uterine cancer, oesophageal cancer, kidney cancer, pancreatic cancer, liver cancer and multiple myeloma).^39–42^ We estimated the impact of changes in the distribution of BMI on the incidence of disease by calculating population impact fractions (PIFs).^43^ A PIF is an estimate of the percentage reduction in rate of disease in a population, and is a function of the baseline distribution of a risk factor, the scenario distribution of a risk factor, and the dose-response relationship between the risk factor and a disease. Dose-response relationships were defined by relative risks from meta-analyses of prospective cohort analyses (see Text S1 for references).

For PRIMEtime analyses, we derived baseline incidence, prevalence and case fatality rates for each of the modelled diseases in the UK using incidence, prevalence and mortality data from the Global Burden of Disease (GBD) and the disbayes R package to derive epidemiologically consistent rates of case fatality, which are not explicitly reported in the GBD reports (see details in Text S1).^44^ ^45^ To estimate background trends in disease incidence and case fatality rates, we estimated rates for 2015 (the baseline year) and 2005 and allowed a linear annual progression between these years to continue into the forecast range for ten years at which point incidence and case fatality rates remain constant.

### Quality adjusted life years

From simulation of the obesity-related diseases, PRIMEtime estimates the impact of the SDIL on population mortality and life expectancy. We determined quality-adjusted life years (QALYs) by adjusting the predicted years of life lived by disease-specific utility weights (see Text S1) which reflect quality of life associated with diseases at each age and sex.^46^

### Health system costs

We derived health care costs for the 2018-19 budget year and adjusted to the 2015 modelling baseline year using the Consumer Price Index.^47^ National Health Service (NHS) costs of treating disease were derived from Department of Health and Social Care budget allocations to: (a) clinical commissioning groups, which are responsible for hospital and community healthcare services in England; (b) primary care and (c) specialised services, which focus on conditions that are particularly expensive or have a small patient population. From the total costs of treating each modelled disease, we estimated an average cost per prevalent case, by dividing by the average number of prevalent cases in 2018-19. From the total costs of treating all other diseases that were not explicitly modelled in PRIMEtime, we estimated an average cost per person in the population in 2018-19. For dental caries, where outcomes are modelled in units of decayed, missing or filled teeth, rather than population cases, we estimated the average cost of treatment from the total value of NHS dental contract payments, the total Units of Dental Activity that were carried out, and the number of Units of Dental Activity associated with dental clinic fillings and extractions. The GP patient survey on dental care suggests that around 42% of all dental patients opt for private rather than NHS dental care, which is typically associated with higher fees than those charged by a NHS provider.^48^ Thus, our estimates of health care cost impacts for dental care are likely to be conservative.

### Net monetary benefit

Net monetary benefit is a measure favoured by the UK Treasury. It reflects the value of an intervention in monetary terms and can be derived from the net intervention impacts on costs to the health sector and population health, assuming a willingness-to-pay threshold for health.^49^ We determined the net monetary benefit of the SDIL policy from PRIMEtime estimates of the lifetime health (QALY) and cost impacts. A QALY was valued at £60,000 as recommended by UK Treasury,^50^ and health care costs were fixed at 2015 prices. Both QALYs and costs were discounted using declining long term discount rates, as recommended by UK Treasury, to capture both social time preferences and intergenerational transfer of wealth: 3.5% costs and 1.5% health (0 to 30 years), 3% costs and 1.29% health (31-75 years) and 2.5% costs and 1.07% health (76-125 years).^51^

### PRIMEtime simulation

We ran both open- and closed-cohort simulations with the PRIMEtime model. To report changes in prevalence of overweight/obesity and rates of disease, we ran the PRIMEtime model until to 2055 while continuing to add new birth cohorts to the simulation, so that the denominator for estimating rates would remain representative of the whole population (open cohort simulation). Estimates of future births in the UK were based on population projections from the Office for National Statistics.^52^ To estimate lifetime impact of the SDIL on QALYs and costs to the health sector, we ran the PRIMEtime model until everyone in the 2015 UK population had died (a closed cohort analysis).

### Uncertainty Analyses

We estimated 95% uncertainty intervals around all model outputs, by repeating the simulations 3000 times, iteratively drawing from uncertainty distributions around PRIMEtime inputs, including the SDIL effect on sugar, relative risks of obesity-related diseases, the sugar-DMFT dose-response relationship, disease utility weights and health care costs. For health care costs, we were only able to derive point estimates for unit costs, so for uncertainty analyses we assumed that the cost followed a lognormal distribution with the point estimate as mean and standard deviation 20% of the point estimate. Contribution of these parameters to the uncertainty in model outputs was assessed in univariate analyses and displayed in a tornado diagram.^53^ The total number of iterations (3000) was sufficient to report stable outcomes to three significant figures.

### Sensitivity analyses

We examined the sensitivity of the modelled estimates of QALYs, costs and net monetary benefit to a number of factors, including the addition of diseases not included in the study protocol, but with evidence of an association with obesity, alternative discount rates, variations in background trends in BMI and disease rates, and the rate at which change in obesity is assumed to have an impact on disease incidence (Table 1). We also evaluated net monetary benefit at a range of willingness-to-pay values for the QALY, ranging from zero to double the UK Treasury rate.

**Table 1.** Sensitivity analyses.

| **Sensitivity analysis** | **Base case scenario** | **Sensitivity scenario** |
| --- | --- | --- |
| Added diseases | Ischaemic heart disease, ischaemic stroke, intracerebral haemorrhage, hypertensive heart disease atrial fibrillation/flutter, diabetes mellitus type 2, colorectal cancer, post-menopausal breast cancer, uterine cancer, oesophageal cancer, kidney cancer, pancreatic cancer, liver cancer and multiple myeloma | As for base case scenario + asthma, low back pain, osteoarthritis of the hip and knee, depression, and gallbladder and biliary diseases.<sup>54-60</sup> |
| BMI linear trend | Non-linear background trend in BMI (model predicts that slowing rates of increase in BMI will ultimately reach an asymptote – see Figure S2 in Text S1).<sup>37</sup> | Linear trend (model predicts that slowing rates of increase in BMI will peak and then decrease – see Figure S2 in Text S1).<sup>37</sup> |
| BMI no trend | Non-linear background trend in BMI (model predicts that slowing rates of increase in BMI will ultimately reach an asymptote – see Figure S2 in Text S1).<sup>37</sup> | No background trend in BMI, i.e. prevalence of overweight and obesity remain stable at 2015 levels. |
| No disease trends | Trends in disease incidence and case fatality applied for 10 years and then remain stable. | No background trends in disease rates; rates remain stable at 2015 levels. |
| No effect lags | Linear increase to full effect of BMI changes on disease incidence at 5 years for cardiovascular and metabolic diseases and 20 years for cancers. | Immediate effect of BMI changes on disease incidence. |
| NICE discount rates | UK Treasury tiered rates (0-30, 31-75, 76+ years):<sup>51</sup><br>3.5%, 3%, 2.5% for health<br>1.5%, 1.29%, 1.07% for costs | National Institute for Health and Care Excellence (NICE) flat rate of 3.5% for costs and health outcomes.<sup>61</sup> |
| Undiscounted | UK Treasury tiered rates (0-30, 31-75, 76+ years):<sup>51</sup><br>3.5%, 3%, 2.5% for health<br>1.5%, 1.29%, 1.07% for costs | No discounting of costs or health outcomes (i.e. 0%). |

### Patient and Public Involvement

This study did not involve any patients or members of the public.

### Ethics approval

This study used publicly available secondary data and did not involve collecting primary data from human participants. Ethical approval was therefore not required.

## Results

### Overweight and obesity

Assuming the sugar reduction effects of the SDIL are sustained to 2055, the model predicts that the SDIL will lead to a reduction in prevalence of both overweight and obesity in the UK population (*Figure 2*). The combined prevalence of overweight and obesity is reduced by 0.18 percentage points (95% uncertainty interval: 0.059 to 0.31) for males and 0.20 percentage points (0.064 to 0.34) for females. This is equivalent to 685 thousand (220 to 1,160 thousand) fewer males and 760 thousand (245 to 1,290 thousand) fewer females with overweight or obesity.

**Figure 2.**
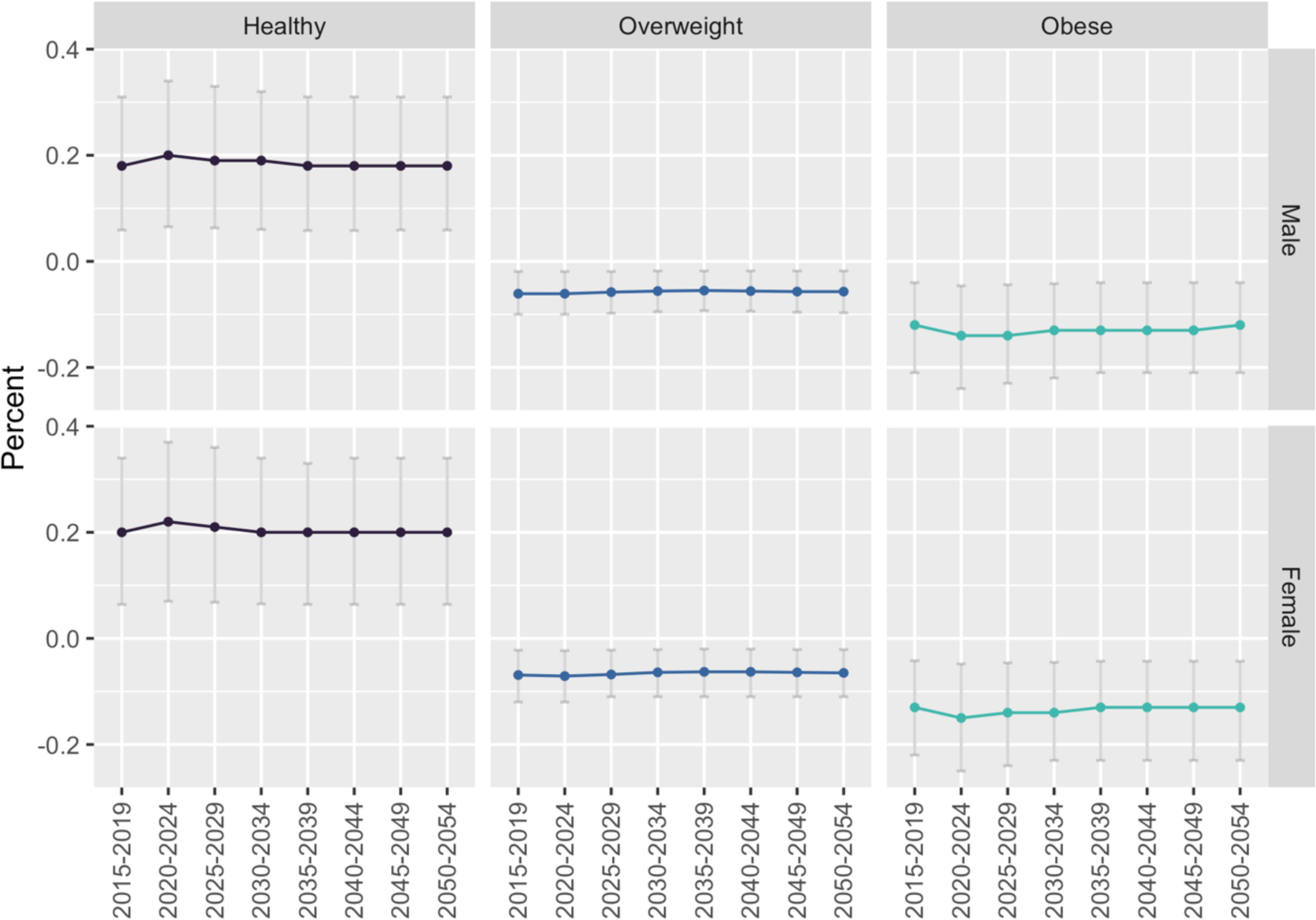
SDIL impact on prevalence of Healthy weight, Overweight and Obesity in the UK population over time

### Obesity-related disease and dental health

Table 2 shows the predicted impact of the modelled reductions in prevalence of overweight and obesity on new cases of obesity-related disease and dental health in the UK population. In the first ten years after implementation of the SDIL, it is estimated there will be 12,000 (3,700 to 20,000) fewer cases of type 2 diabetes, 3,800 (1,200 to 6,700) fewer cases of cardiovascular diseases, 350 (110 to 590) fewer cases of obesity-related cancers, and 270,000 (35,000 to 600,000) fewer dental caries.

**Table 2.**
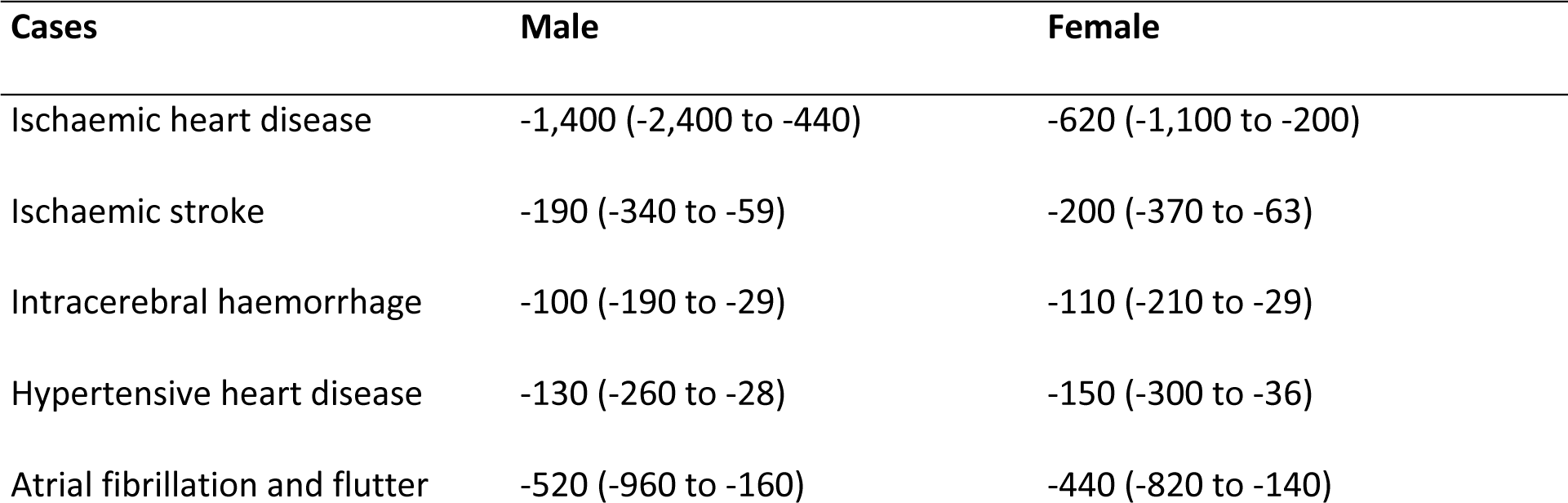
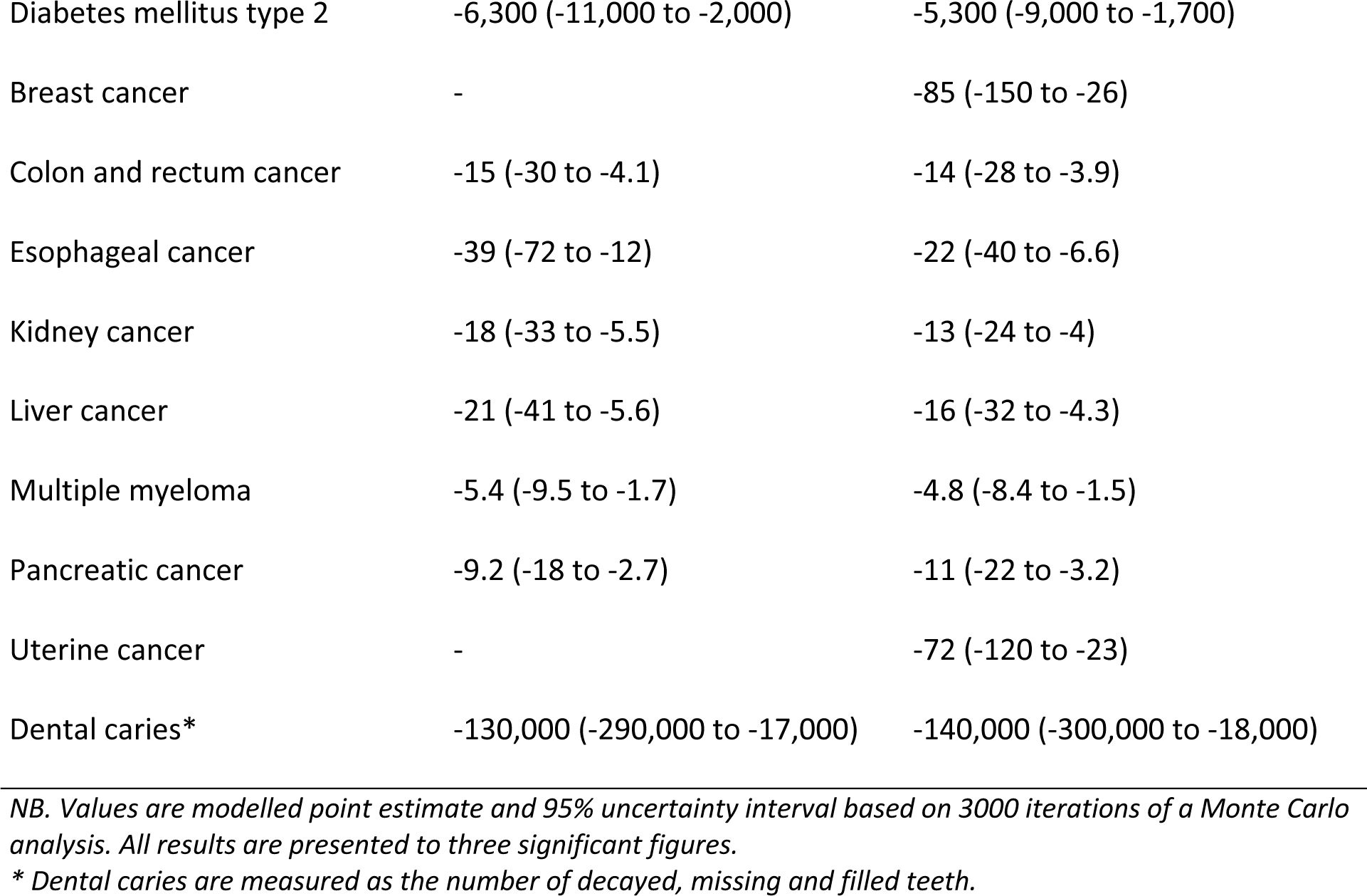
SDIL impact on incident cases of disease and dental caries, in the ten years following SDIL implementation.

### Quality adjusted life years and health care costs

The model predicts that the SDIL will lead to a population health gain of 200,000 QALYs (63,500 to 342,000) over the lifetime of the 2015 UK population (Table 3). It is estimated there will be a £174 million (£53.6 to £320) reduction in costs to the health service for treatment of dental caries and obesity-related diseases, which will be countered by a small £94.6 thousand (£30.6 to £162) increase in costs of treatment for other diseases (e.g. injuries, dementia, etc.) that occur due to additional years of life lived. This leads to a net cost-saving of £174 million (£53.6 to £319). At the UK Treasury value of £60,000 per QALY, the net effect of these modelled lifetime impacts on health and health care costs is a net monetary benefit of £12.2 billion (£3.88 to £20.8). Net monetary benefit is linearly related to the willingness-to-pay value for the QALY: different values can be estimated from Figure S2 in Text S2.

**Table 3.**
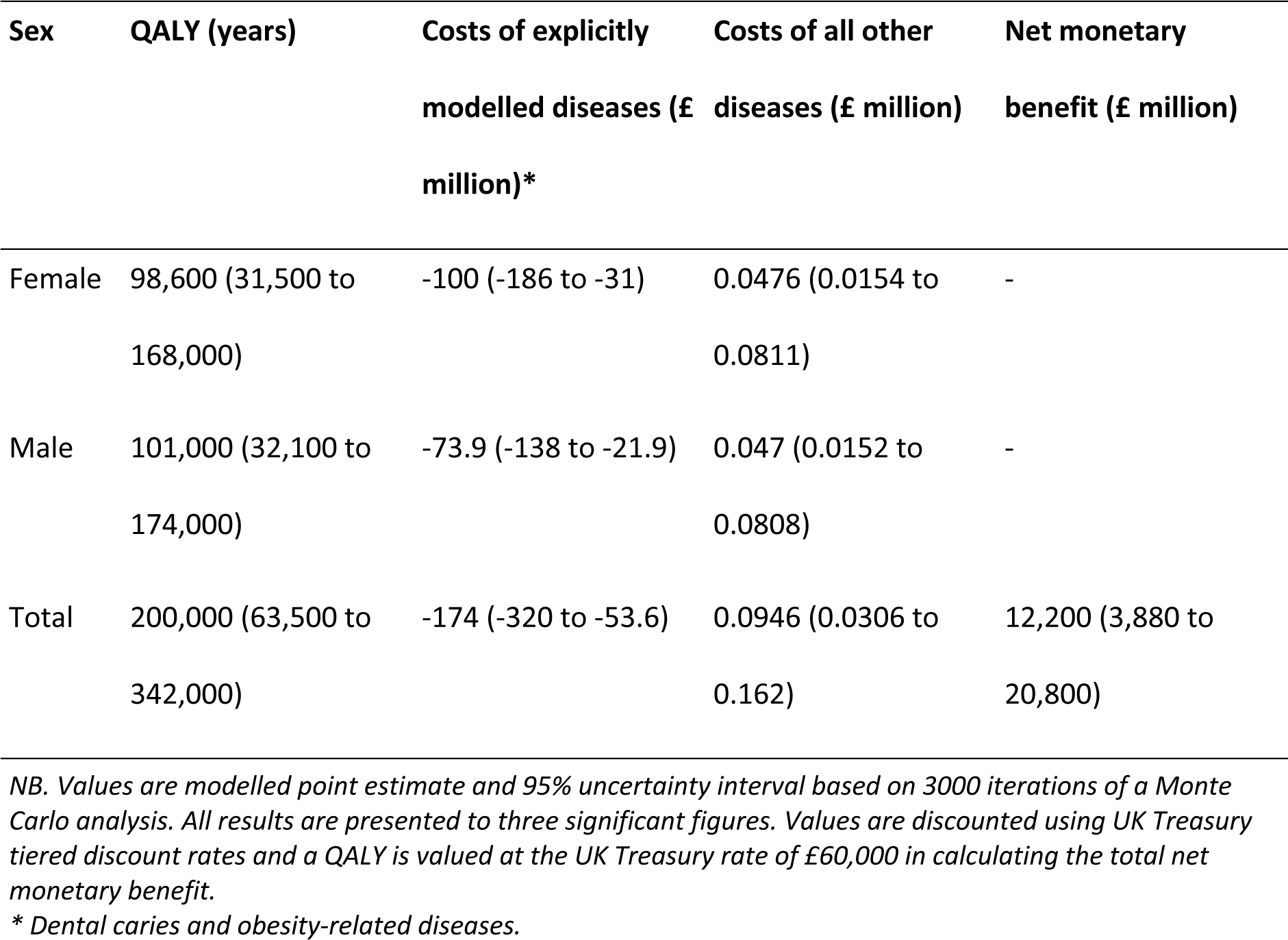
Lifetime population health and health care cost impacts and net monetary benefit of the SDIL.

| Sex | QALY (years) | Costs of explicitly<br>modelled diseases (£<br>million)* | Costs of all other<br>diseases (£ million) | Net monetary<br>benefit (£ million) |
| --- | --- | --- | --- | --- |
| Female | 98,600 (31,500 to<br>168,000) | -100 (-186 to -31) | 0.0476 (0.0154 to<br>0.0811) | - |
| Male | 101,000 (32,100 to<br>174,000) | -73.9 (-138 to -21.9) | 0.047 (0.0152 to<br>0.0808) | - |
| Total | 200,000 (63,500 to<br>342,000) | -174 (-320 to -53.6) | 0.0946 (0.0306 to<br>0.162) | 12,200 (3,880 to<br>20,800) |

*NB. Values are modelled point estimate and 95% uncertainty interval based on 3000 iterations of a Monte Carlo analysis. All results are presented to three significant figures. Values are discounted using UK Treasury tiered discount rates and a QALY is valued at the UK Treasury rate of £60,000 in calculating the total net monetary benefit.*
*\* Dental caries and obesity-related diseases.*

### Uncertainty analyses

Examination of the tornado diagram (Figure 3) shows that the majority of uncertainty in the magnitude of the health gains, net costs and net monetary benefit is due to the wide confidence interval around the effect of the SDIL on sugar in purchased drinks. The uncertainty in the unit costs of disease, which we estimated as a standard deviation equal to 20% of the point-estimate cost, also has sizeable impact on the net health care costs.

**Figure 3.**
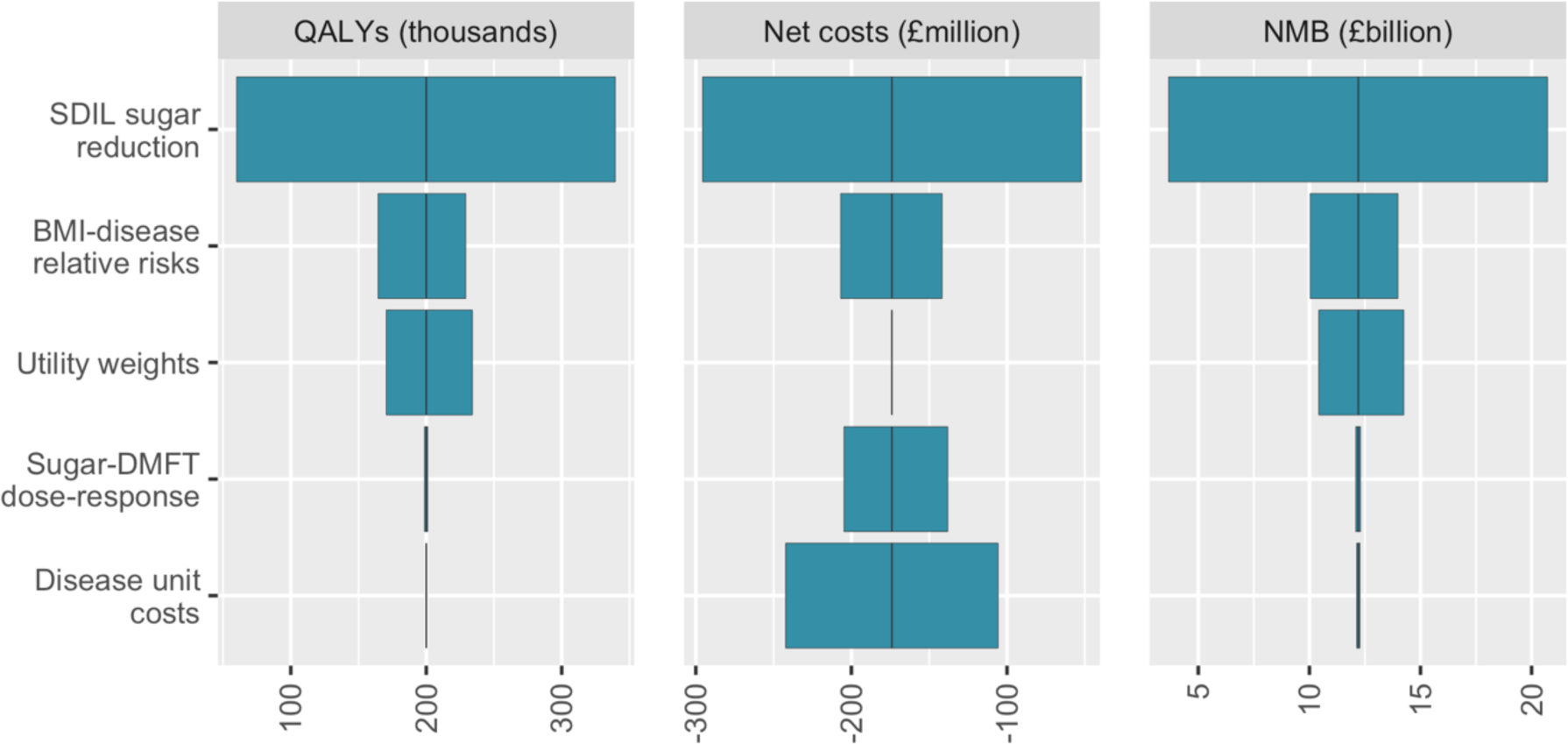
Tornado plots illustrating the contribution of uncertainty around model input parameters to the uncertainty in model outputs – QALYs, net costs and net monetary benefit (NMB).

### Sensitivity analyses

In sensitivity analyses, the health gains, net cost-savings and positive net monetary benefit remained significant under all scenarios evaluated, although the magnitude of the estimates varied (Figure 4).

**Figure 4.**
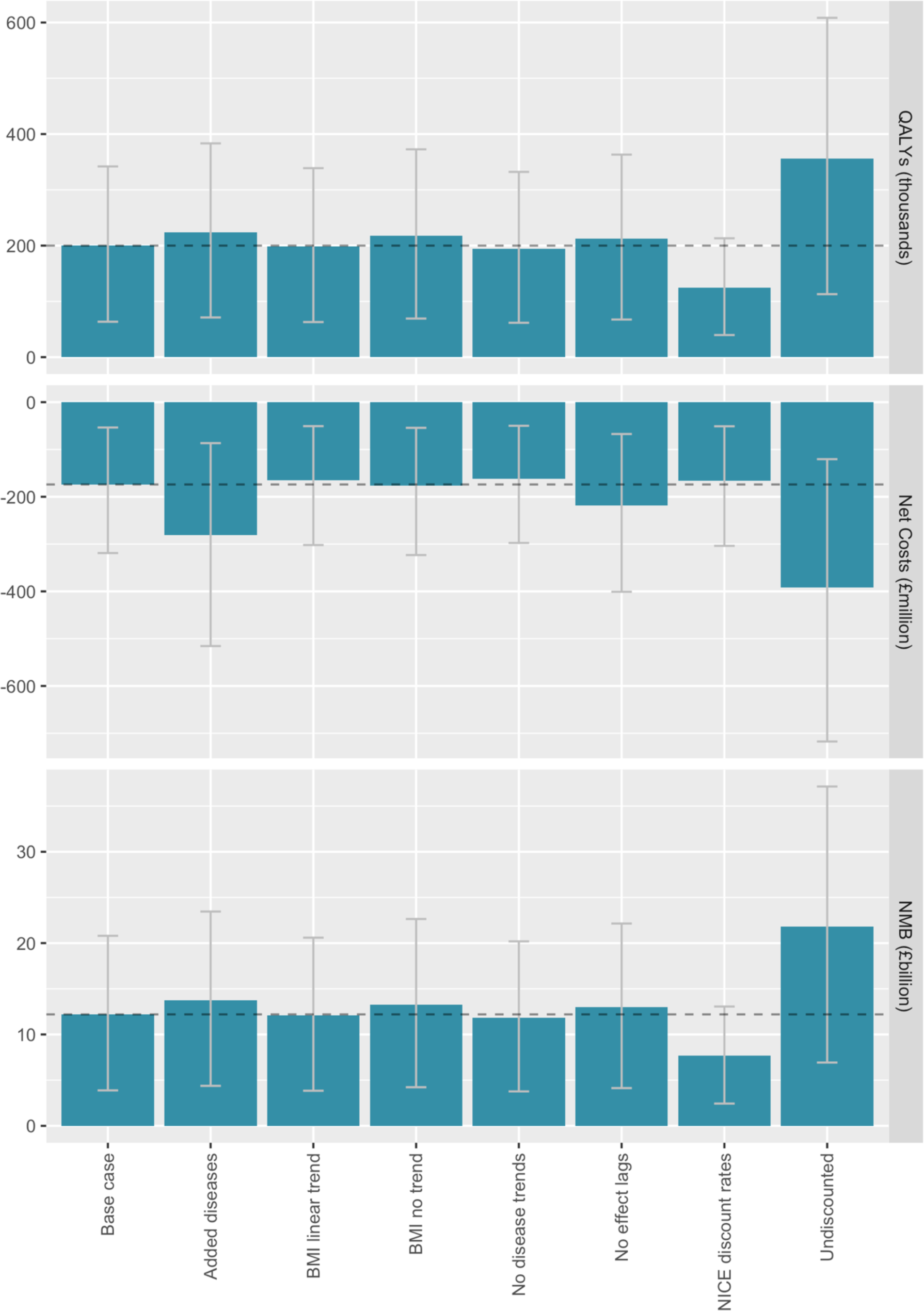
Sensitivity of net monetary benefit to a range of modelling assumptions.

Modelled estimates were most sensitive to the discount rates chosen for analysis. For example, the net monetary benefit of the SDIL was 37% lower if discounted using NICE rates of 3.5% for both QALYS and costs, but 79% higher with no discounting of QALYs or costs. The addition of potential BMI-related changes in asthma, depressive disorders, gallbladder and biliary diseases, low back pain and osteoarthritis of the hip and knee increased cost-savings (+62%) more than it increased QALYs (+12%), which led to a 13% higher net monetary benefit. The choice of background trend model (linear vs non-linear) had relatively little impact on health economic outcomes (<1% change in net monetary benefit), but the net monetary benefit was 8.9% higher when assuming no background trends at all in BMI (see Figure S1 in Text S2 for comparison of the impact on prevalence of overweight and obesity) . Removing the assumed lags in effect of changes in BMI on disease incidence (5 years for cardiovascular disease and 20 years for cancer) increased net monetary benefit by 6.5%, while removing background trends in diseases reduced net monetary benefit by 2.9%.

## Discussion

### Statement of principal findings

We evaluated the population health and health sector impacts of the SDIL, using the PRIMEtime model to simulate the likely impact on obesity, disease and health care costs from the observed effects on sugar purchased in drinks for in-home consumption. The model predicts that the reductions in sugar will reduce prevalence of overweight and obesity in the UK, preventing type 2 diabetes, cardiovascular disease and obesity-related cancers, and it will improve dental health. For the current UK population, it is estimated the SDIL will add 200,000 QALYs (63,500 to 342,000) over their lifetime and avert £174 million (£53.6 to £319) in their costs of health care, producing a net monetary benefit of £12.2 billion (£3.88 to £20.8) for the health system. There are relatively wide uncertainty intervals around the model predictions, which chiefly reflect the wide uncertainty in the effect of the SDIL on sugar purchased in drinks (-8.0 g/household/week [95% confidence interval: -13.6 to -2.4]).^16^ Nevertheless, the beneifical impacts of the SDIL are robust to variations in modelling assumptions around background trends in BMI and disease rates, lags in the effect of the SDIL on disease, and discount rates.

### Comparison with other literature

Our results are consistent with previous modelling studies, which have predicted health and monetary benefits of a tax targeting sugar-sweetened beverages in the UK.^12^ ^13^ Since these studies preceded implementation of the SDIL, they relied on price elasticity data to estimate the likely consumer response to a hypothetical tax on sugar-sweetened beverages. Such studies cannot directly capture potential supply-side responses, such as reformulation, or consumer responses to the signalling of health concerns potentially associated with a government initiating an intervention targeting sugar-sweetened beverages.^62^ By drawing on real-world evaluation of the SDIL, our modelling study provides stronger evidence of the likely health and health sector benefits of implementing a tax targeting sugar-sweetened beverages in the UK.

As of December 2021, 40 countries are reported to have announced or implemented some form of sugar-sweetened beverage tax.^14^ There is a growing body of evidence demonstrating the effectiveness of taxes in reducing purchasing or consumption of sugar-sweetened beverages,^62^ but to date only in Mexico has this evidence from real-world evaluation been used to examine the future consequences for population health and health care expenditure. Like our study, modelling of the sugar-sweetened beverage tax implemented in Mexico^55^ estimated that the tax would be a cost-effective intervention for improving population health from a health care perspective.^63^ There are differences in the design of the taxes between the two countries: Mexico implemented a 1 peso per litre excise tax, which raised the retail price of drinks; whereas the UK implemented a tiered levy on drinks, as an incentive for drink manufacturers to reduce the sugar content of drinks. But overall, the results of the modelling analyses were relatively similar: predicting small but significant reductions in obesity prevalence (<1%) leading to large reductions in cases of diabetes, cardiovascular disease and cancers, and reduced health care expenditure.

### Strengths and weaknesses of the study

A key uncertainty in modelling the population health impacts of sugar-sweetened beverage taxes is the pathway by which the change in drink consumption or sugar-content of drinks influences disease. The evaluation of the UK SDIL found a reduction in sugar purchased in drinks.^16^ In our modelling analyses, we estimated the equivalent reduction in calories, and simulated effects of reduced energy intake on BMI and obesity-related disease. There is a wealth of observational and trial evidence supporting the causal relationship between the intake of sugar and sugar-sweetened beverages, and obesity.^9^ ^64^ But there is also a growing body of evidence, including lab studies and prospective cohort studies, suggesting that the metabolism of sugars in sugar-sweetened beverages may have cardio- metabolic effects that are over and above the impacts on energy intake that we have accounted for in our modelling.^65^ By restricting our cardiovascular disease and cancer modelling to the direct calorific impact on body weight, we may have underestimated the full impact of the SDIL.

We also did not evaluate the effect of any change in consumption of non-sugar sweeteners that may have occurred in the reformulation of drink products with the SDIL. Randomised controlled trials suggest that people consuming non-sugar sweeteners as a replacement for sugars have a lower body weight or BMI at the end of the trial, but longer-term prospective cohort studies suggest there be an increased risk of diabetes and cardiovascular disease with consumption of non-sugar sweeteners.^66^ It is possible these observed associations with harmful effects are due to reverse causation and/or residual confounding. We did not model the effects, if any, of possible increases in consumption of non-sugar sweeteners on population health due to the SDIL.

The interrupted time series analysis of the SDIL was based on household-level purchasing data, hence we were not able to determine how the changes in household sugar purchasing may have impacted differently on individuals within the household (e.g. adults vs children). Additionally, the interurupted time series analysis only included data on products brought into the home.^36^ Out-of-home purchases account for around 10-12% of expenditure on cold non-alcohol beverages in the UK.^67^ If the SDIL has a similar impact on sugar in drinks purchases out-of-home then the SDIL may have had a larger impact on health than we have modelled here, but further work is needed to understand the impacts of the SDIL on out-of-home purchases.

Additionally, we do not know the impact of product wastage within the home. The reduction in sugar that we have modelled is a net effect of changing sugar content and changing purchase volume of taxed and untaxed products. Analyses of UK household waste in 2012 estimated volumetric wastage proportions of 5.2% for bottled water, 7.2% for carbonated soft drink, 8.6% for squash, 12.0% for fruit juices and smoothies and 7.0% for milk.^68^ This suggests that waste may vary by drink type, but the net waste effect due to changing proportions of different product purchasing was not estimated in the time series analyses of the SDIL effect. Further, we do not know if product wastage is influenced by possible taste changes due to product reformulation.

### Implications

Sugar-sweetened beverage taxes are among the suite of “best buy” interventions recommended by the World Health Organization (WHO) for addressing childhood obesity and preventing non-communicable diseases.^69^ ^70^ From an economic perspective, the costs of treating dental caries and obesity-related diseases that stem from drinking sugar-sweetened beverages are a negative externality, and taxes are a potential way of internalising these costs. Our modelling of the UK SDIL and the modelling of the sugar-sweetened beverage tax in Mexico indicate that these interventions are likely to both improve population health and reduce health sector expenditure.

The WHO recommends limiting free sugar consumption to a maximum of 10% of energy intake.^71^ The high levels of potential harm, low nutritional benefit and discretionary nature of sugar-sweetened beverages make them an ideal target for reducing free sugar intake. In the UK, however, the SDIL on its own will not be enough to reduce free sugar consumption to the levels recommended by the WHO. In the UK, non-alcoholic beverages account for a large proportion of free sugar intake, particularly in teenagers (34%), but across all ages the majority of free sugars are consumed in foods such as cereal and cereal products (e.g. cakes, pastries),discretionary sugars (e.g. in tea and coffee), preserves and sweet spreads, confectionery and dairy products (e.g. yoghurt and dairy desserts).^72^ Widening the remit of food taxes to include free sugar from all sources could be a useful tool to prompt both reformulation and reduce purchasing of high sugar foods.^73^ ^74^ Such a tax was proposed in the National Food Strategy review commissioned by the UK Government.^75^ Estimates suggest this tax may reduce [free] sugar consumption by as much as 4-10g/person/day,^76^ which is substantially more than the sugar reduction associated with the SDIL (approximately 0.48 g/person/day),^16^ but further modelling of the National Food Strategy tax scenarios is needed to fully account for changes in demand across the food system. Studies from Australia and New Zealand suggest that there are likely to be population health benefits from combining food and drink taxes and/or subsidies, such as a tax on sugar-sweetened beverages and a subsidy on fruits and vegetables.^77^ ^78^ But further work exploring the public acceptability of fiscal policies in the food system, and working with the public to design food tax and subsidy scenarios, may also help to build trust and political support for new interventions in the UK.

### Unanswered questions and future research

In this study we examined impacts of the SDIL on population health and costs of health care, but there are also likely to be broader societal impacts stemming from reductions in obesity-related diseases, such as increased productivity in working-age adults and reduced costs of social care at older ages. We determined net monetary benefit of the SDIL from the averted health care costs and by applying a value of statistical life to the QALY gain, but we did not include costs of delivering the intervention. However, in ongoing macroeconomic work we are examining a much wide range of impacts on UK Treasury, industry and consumers (e.g. changes in revenue, employment, GDP and household spending) and in forthcoming assessment we will be presenting a wider range of both health and economic indicators. Additionally, in ongoing work we are exploring the socio-economic implications of the SDIL for population health.

## Conclusion

The UK SDIL is a tiered levy designed to encourage drink manufacturers to reduce sugar content. Analysis at one year after implementation in April 2018 found that it had reduced sugar in drinks purchased for home consumption by 8.0 g/household/week (95% confidence interval: 2.4 to 13.6). Population health modelling suggests that these changes in sugar consumption, if sustained, will reduce prevalence of overweight/obesity and related diseases and improve dental health in the UK. Health economic analysis indicates that over the lifetime of the current UK population the SDIL could add 200,000 quality-adjusted life years (63,500 to 342,000) and avert £174 million (£53.6 to £319) in health care costs, leading to a net monetary benefit of £12.2 billion (£3.88 to £20.8) for the health sector. This study provides further evidence that sugar-sweetened beverage taxes have the potential to achieve meaningful improvements in population health and reduce health sector spending.

## Contributor and guarantor information

PS acts as guarantor for this manuscript. Contributions to development of the manuscript are as follows: Conception of idea – LJC, RS, HR, MR, AB, MW, PS; Study design – LJC, CL, OM, AB, MW, PS; Grant application – RS, SC, HR, MR, OM, AB, JA, MW, PS; Literature review – LJC, PS; Development of methods – LJC, CL, PS; Data management – LJC, CL; Data analysis – LJC; Data interpretation – LJC, CL, RS, SC, OM, HTJ, MK-B, JA, MW, PS; First draft of manuscript – LJC; Comments on drafts – all authors. The corresponding author attests that all listed authors meet authorship criteria and that no others meeting the criteria have been omitted.

## Copyright / license to publish

The Corresponding Author has the right to grant on behalf of all authors and does grant on behalf of all authors, a worldwide licence to the Publishers and its licensees in perpetuity, in all forms, formats and media (whether known now or created in the future), to i) publish, reproduce, distribute, display and store the Contribution, ii) translate the Contribution into other languages, create adaptations, reprints, include within collections and create summaries, extracts and/or, abstracts of the Contribution, iii) create any other derivative work(s) based on the Contribution, iv) to exploit all subsidiary rights in the Contribution, v) the inclusion of electronic links from the Contribution to third party material where-ever it may be located; and, vi) licence any third party to do any or all of the above.

## Data sharing statement

All data used to support the PRIMEtime model and the analyses conducted in this paper are available in the public domain, some with licensed agreements from relevant data archives (e.g. the UK Data Archive https://www.data-archive.ac.uk/).

## Transparency declaration

The contributing author and guarantor (PS) affirms that the manuscript is an honest, accurate, and transparent account of the study being reported; that no important aspects of the study have been omitted; and that any discrepancies from the study as originally planned have been explained.

## Data Availability

All data used to support the PRIMEtime model and the analyses conducted in this paper are available in the public domain, some with licensed agreements from relevant data archives (the UK Data Archive https://www.data-archive.ac.uk/).

## Text S1 – PRIMEtime data inputs

### 1. Population numbers

**Table S1.**
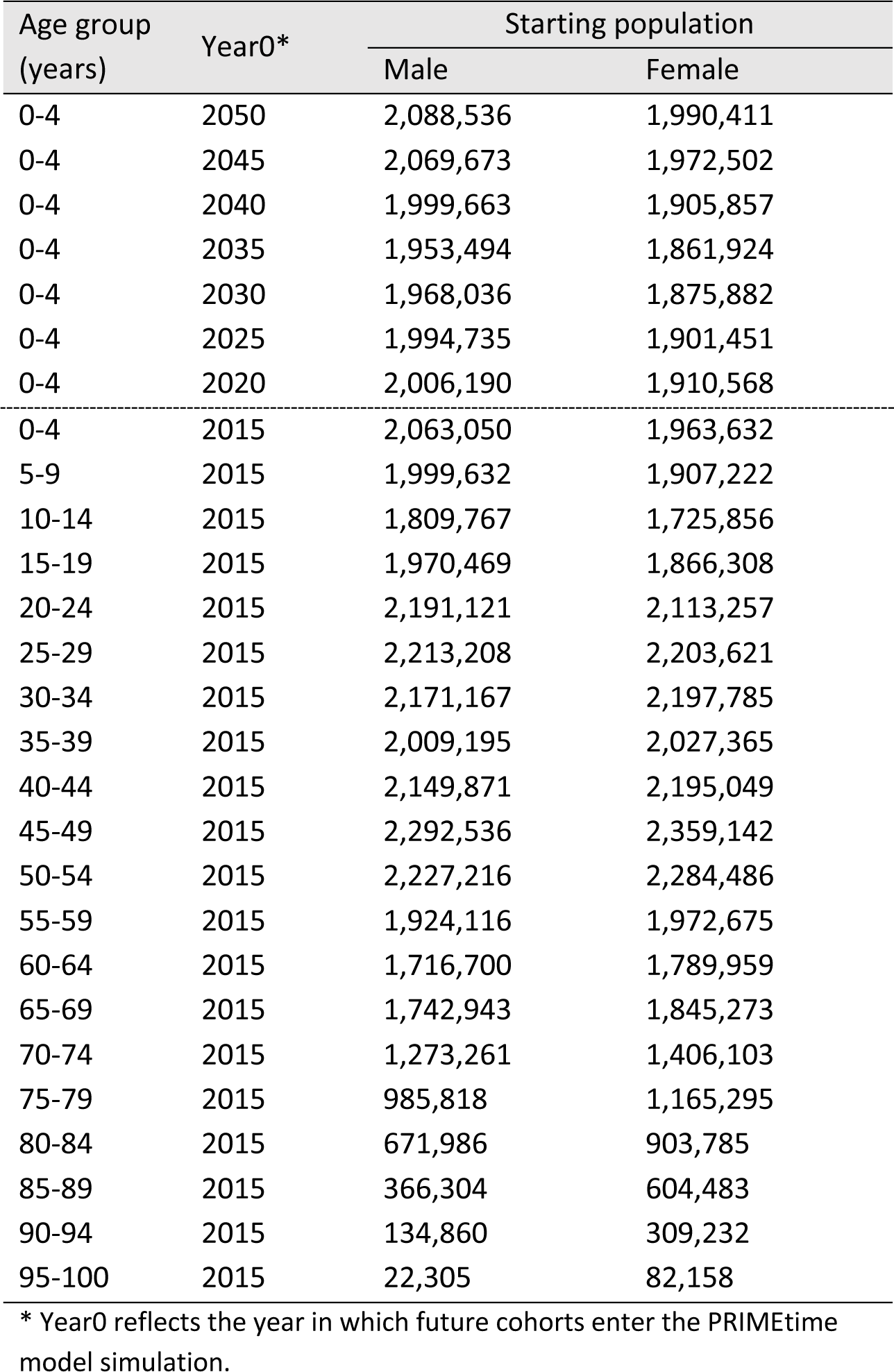
UK population by age and sex from the Human Mortality Database^1^ (all ages in 2015) and population projections from the Office for National Statistics^2^ (future 0-4 year olds)

### 2. All-cause mortality rates

**Figure S1.**
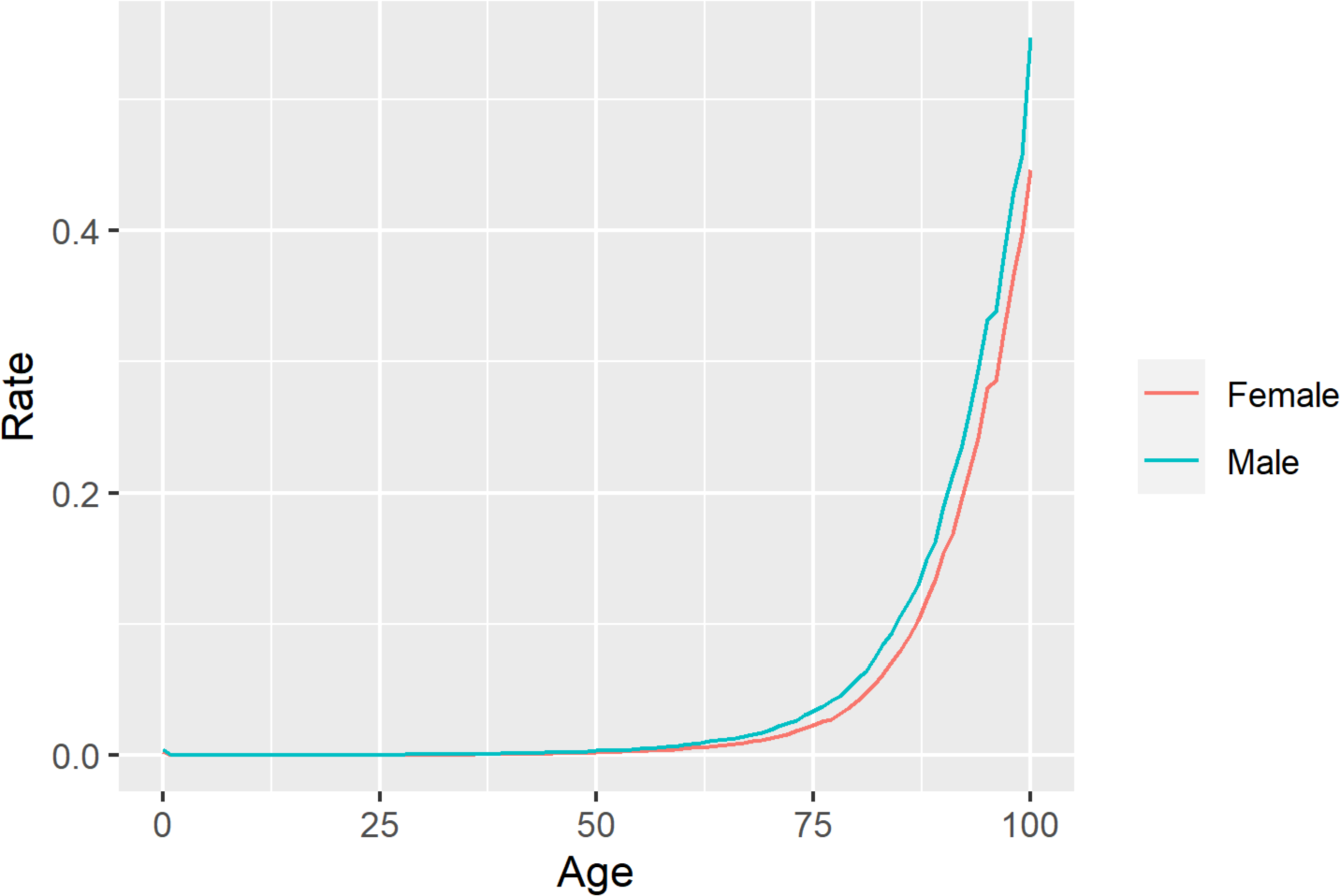
UK all-cause mortality, by single year of age and sex, from the Human Mortality Database.^1^

### 3. Background trends in overweight and obesity

**Figure S2.**
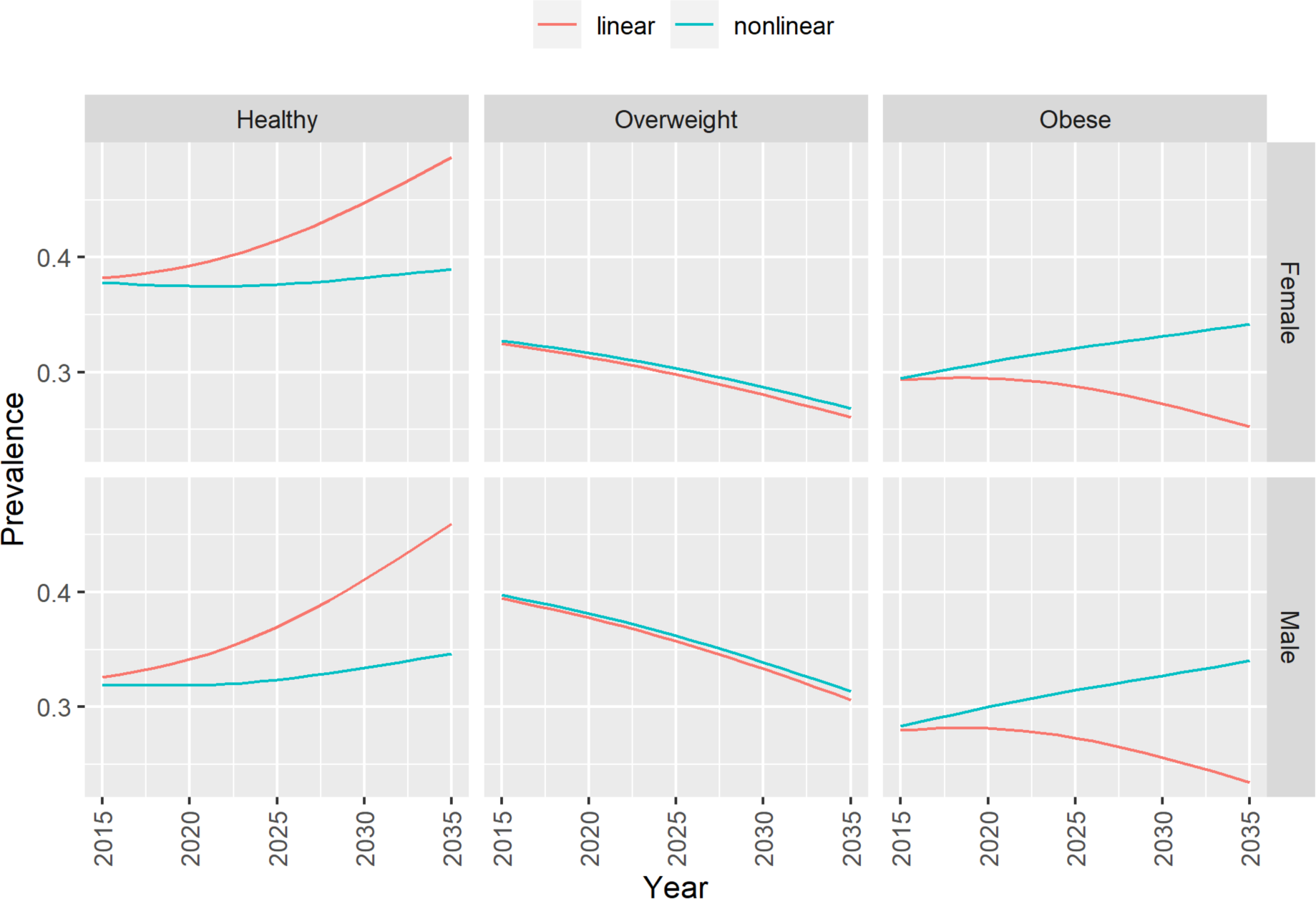
Predicted trends in overweight and obesity with non-linear (base case) and linear (sensitivity) models from Cobiac et al.^3^

### 4. Dental caries rates

**Figure S3.**
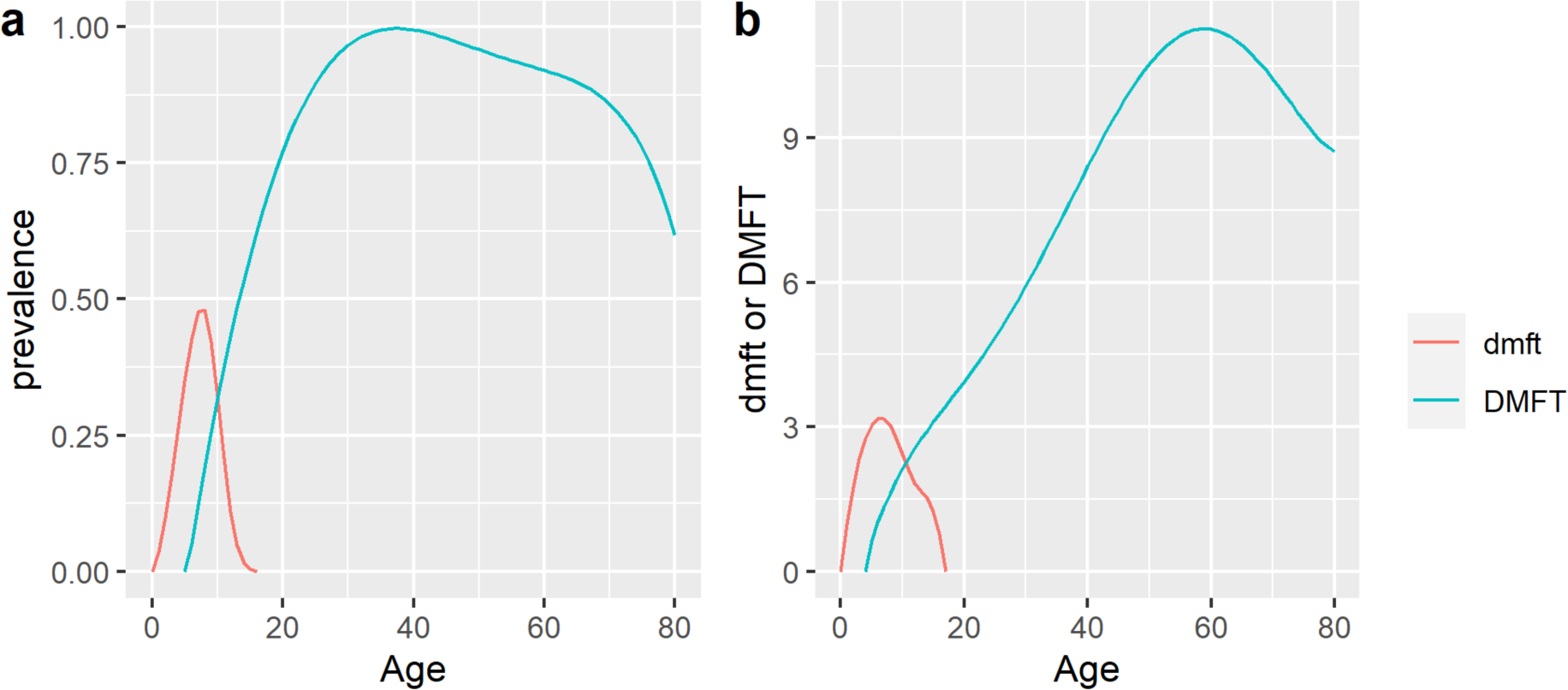
(a) prevalence of and (b) mean number of decayed, missing and filled teeth that are deciduous (dmft) and permanent (DMFT); derived from data collected in the Child and Adult Dental Health Surveys,^4, 5^ adjusted for congenital tooth absence,^6^ and causes of tooth extraction.^7, 8^

### 5. Disease incidence rates

**Figure S4.**
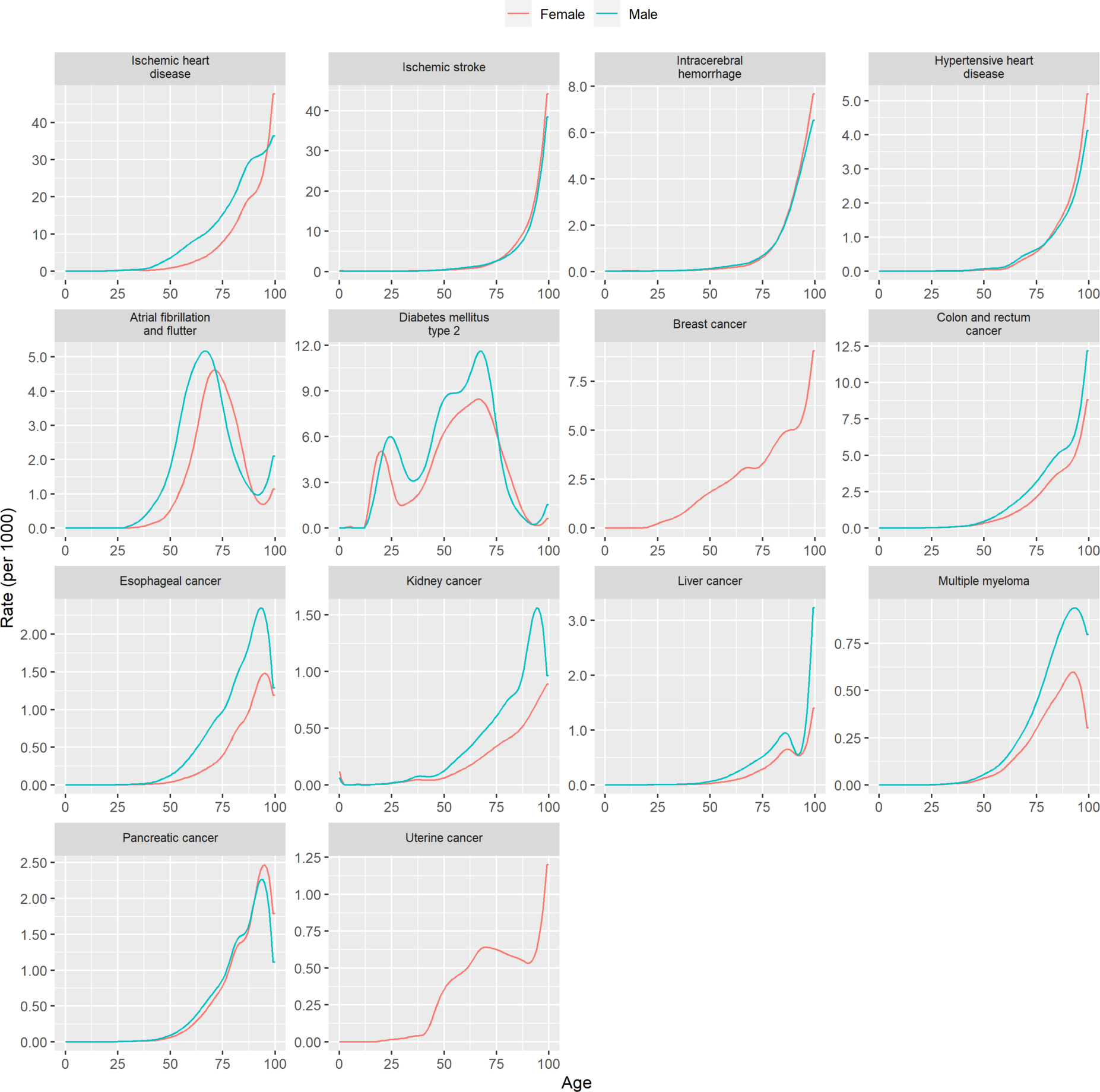
Incidence rates, derived from Global Burden of Disease^9^ estimates using disbayes.^10^ (NB. Graphs are presented on different scales to show detail, but in some cases this may give an exaggerated appearance of variability across ages.

### 6. Disease case fatality rates

**Figure S5.**
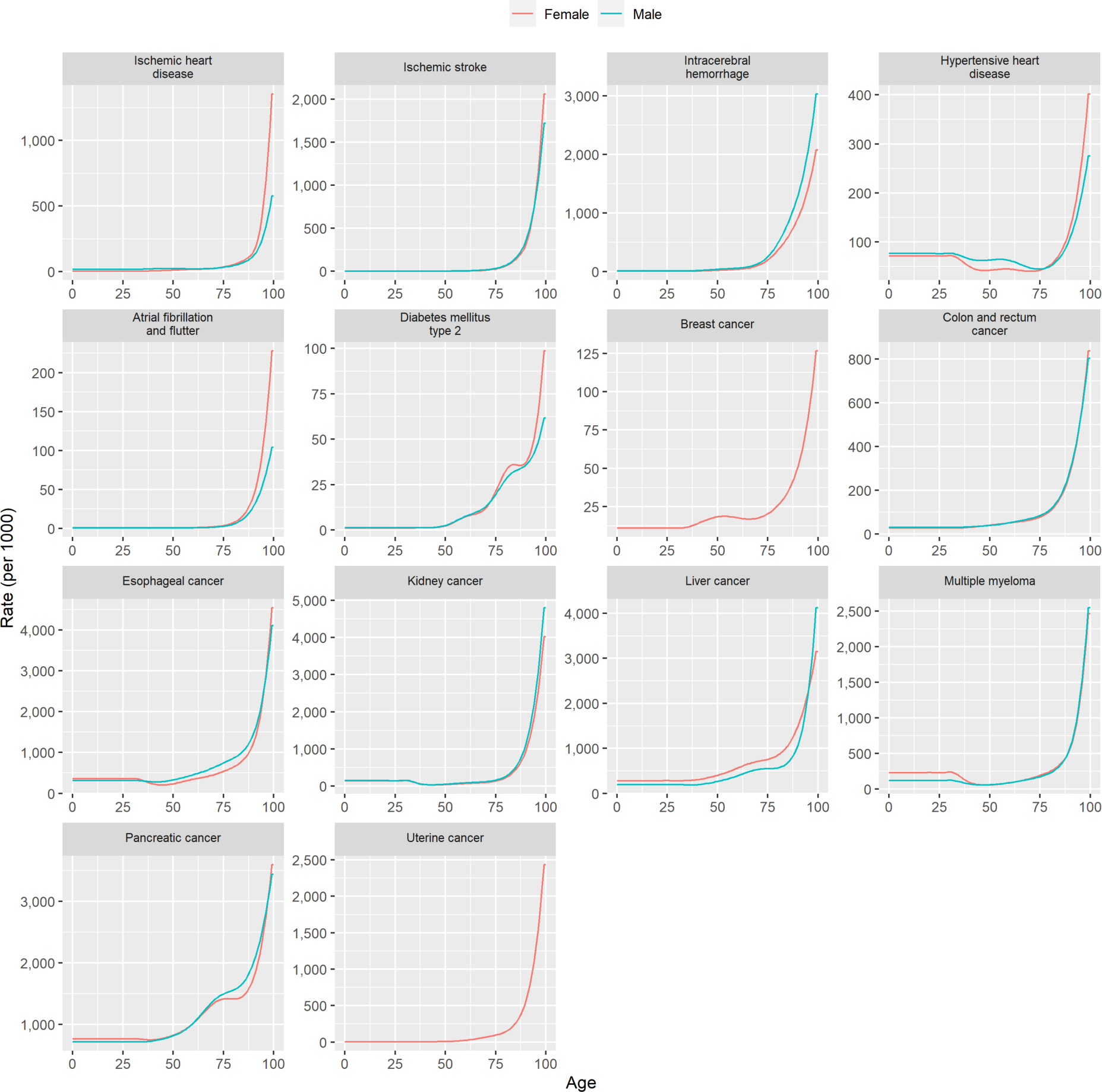
Case fatality rates, derived from Global Burden of Disease^9^ estimates using disbayes.^10^

### 7. Disease prevalence

**Figure S6.**
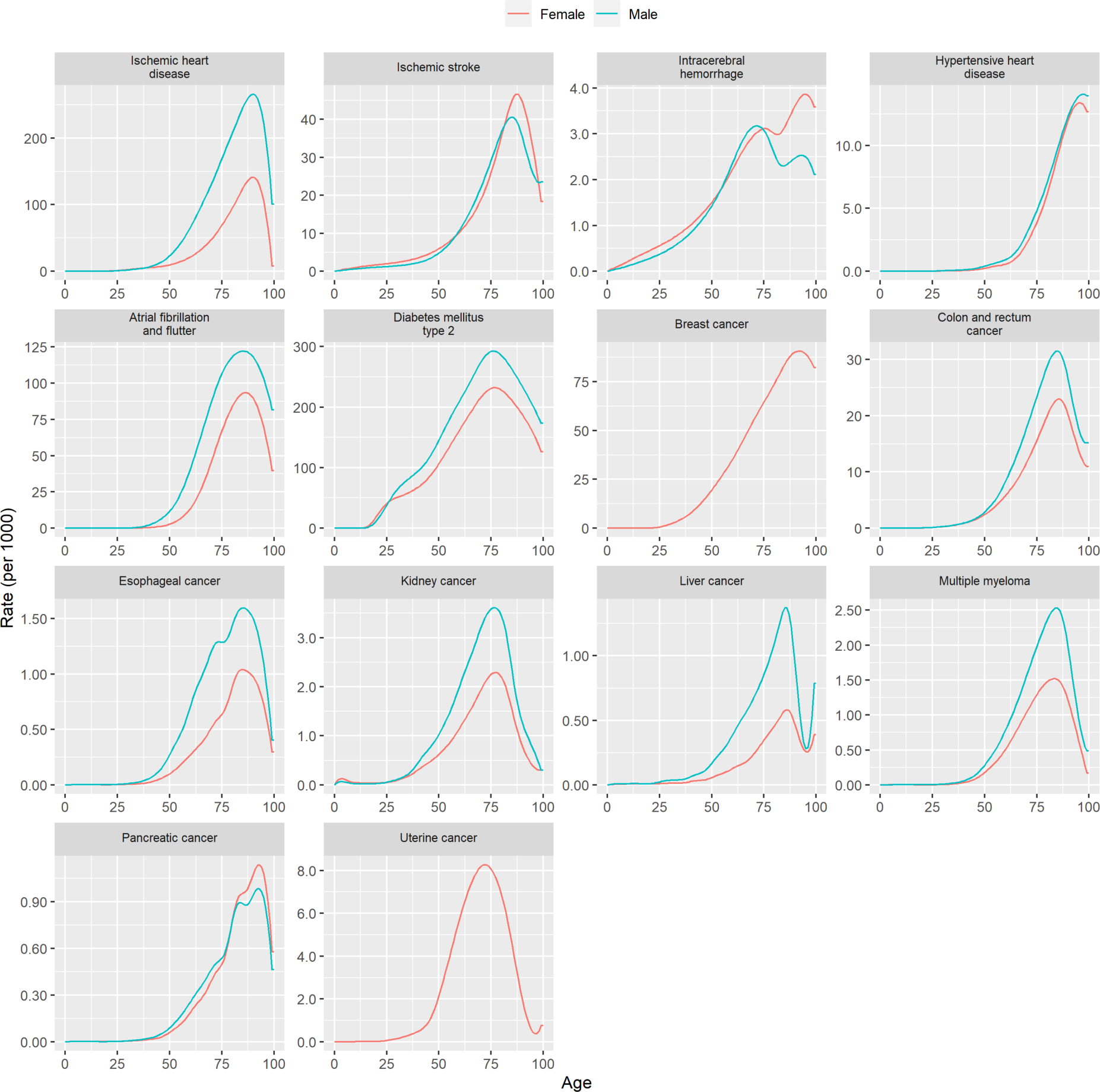
Starting prevalence rates, derived from Global Burden of Disease^9^ estimates using disbayes.^10^

### 8. Background trends in disease incidence and case fatality

**Table S2.**
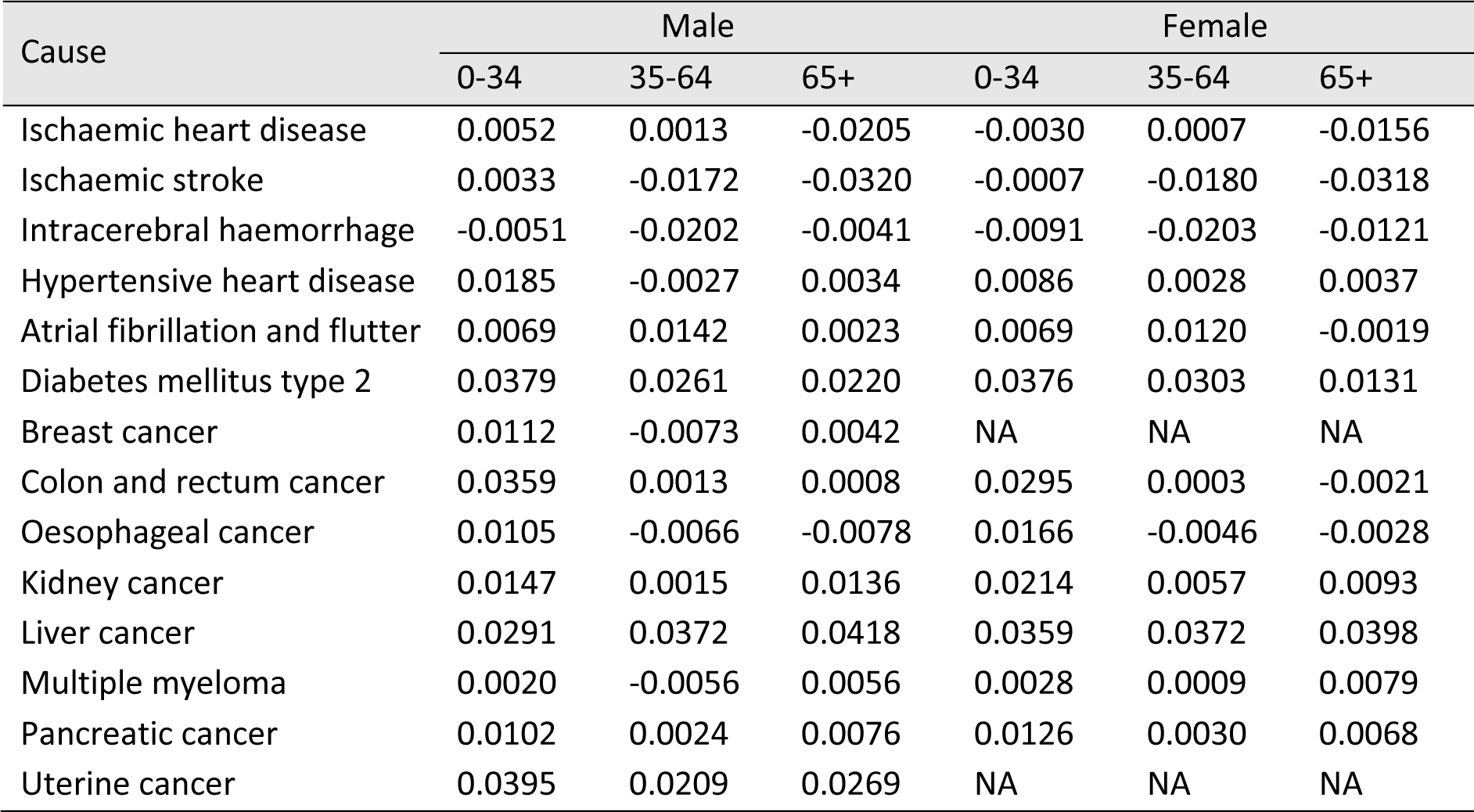
Annual trends in incidence rates, by sex and age group.

**Table S3.**
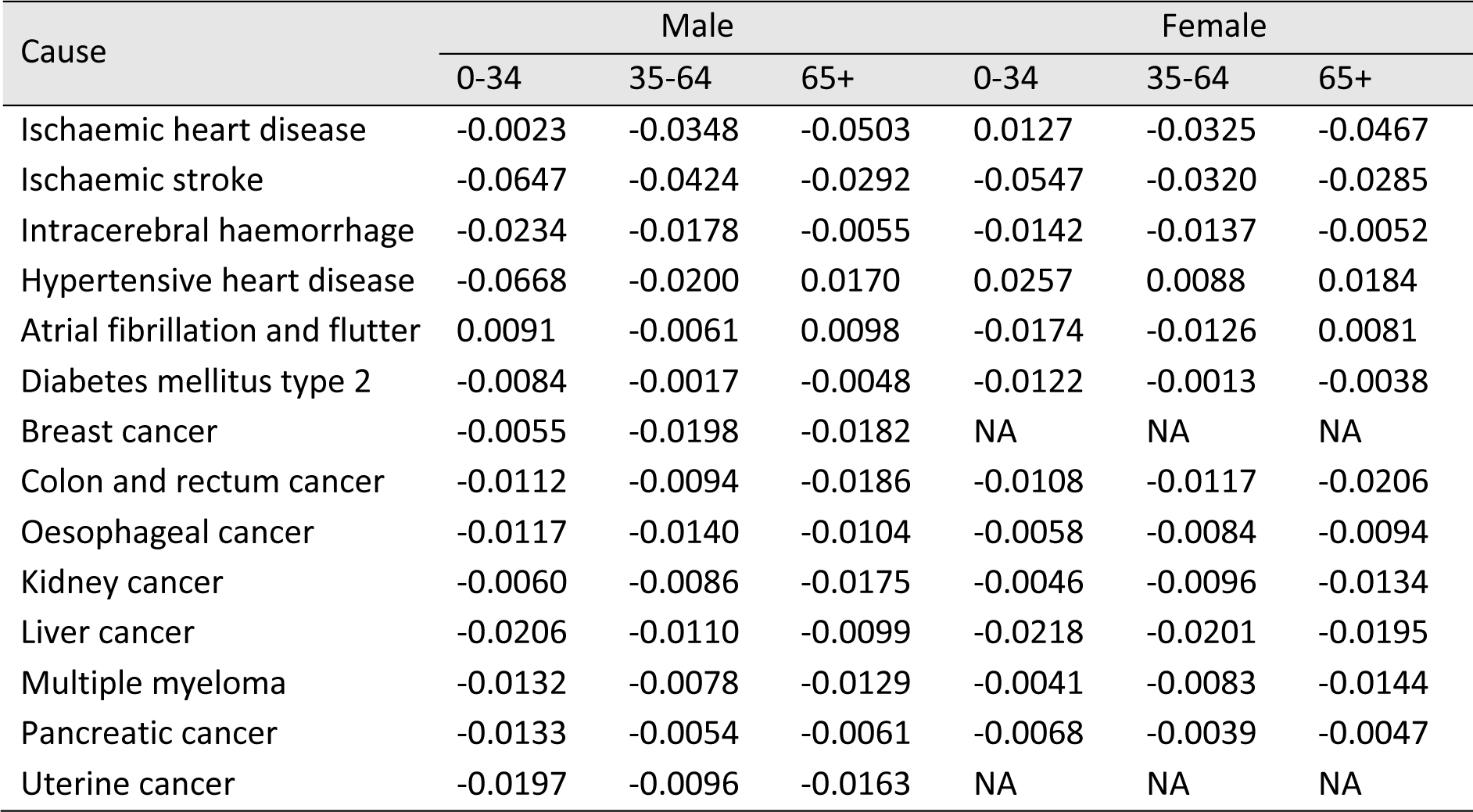
Annual trends in case fatality rates, by sex and age group.

### 9. Relative risks of disease

**Table S4.**
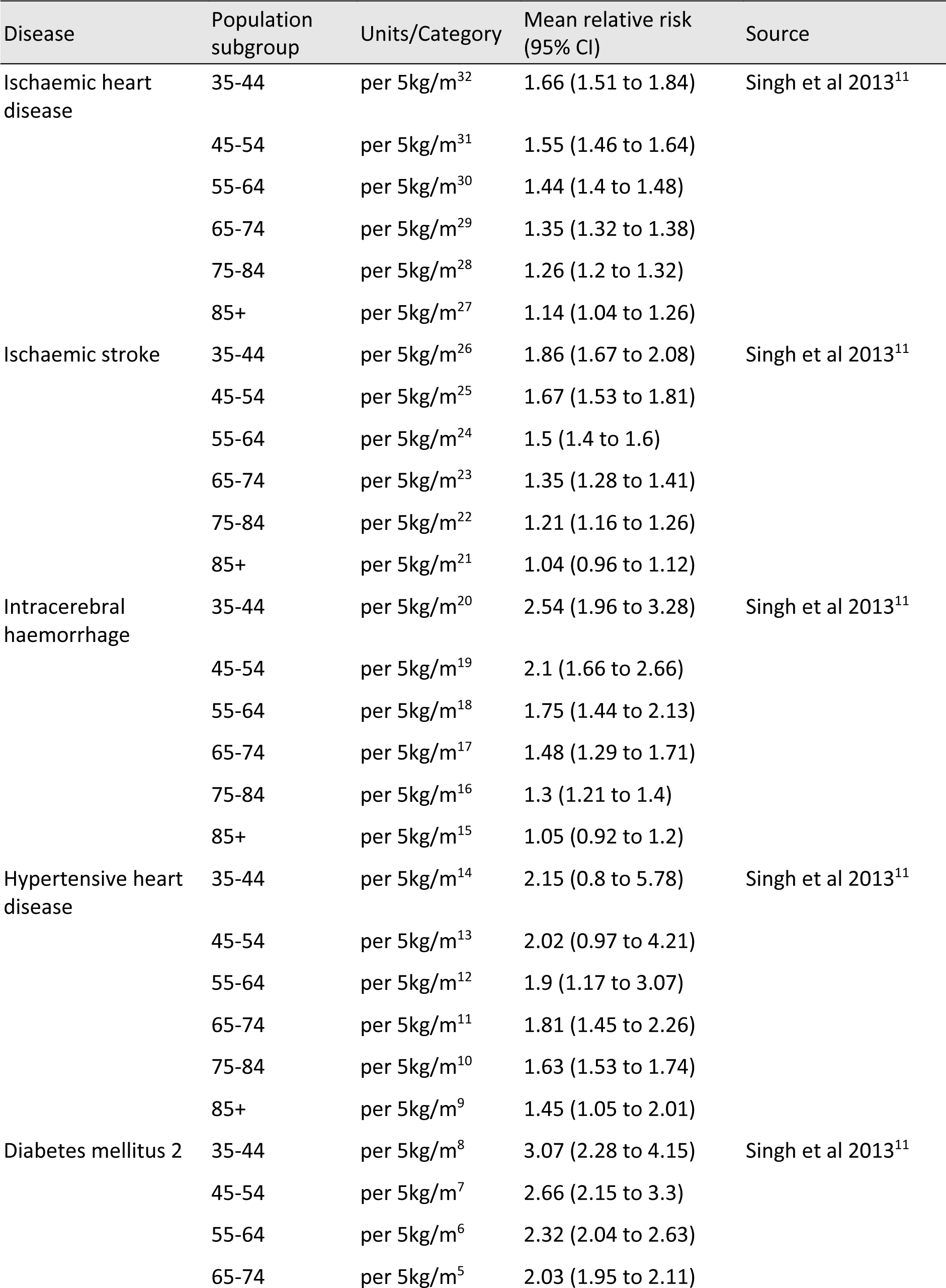
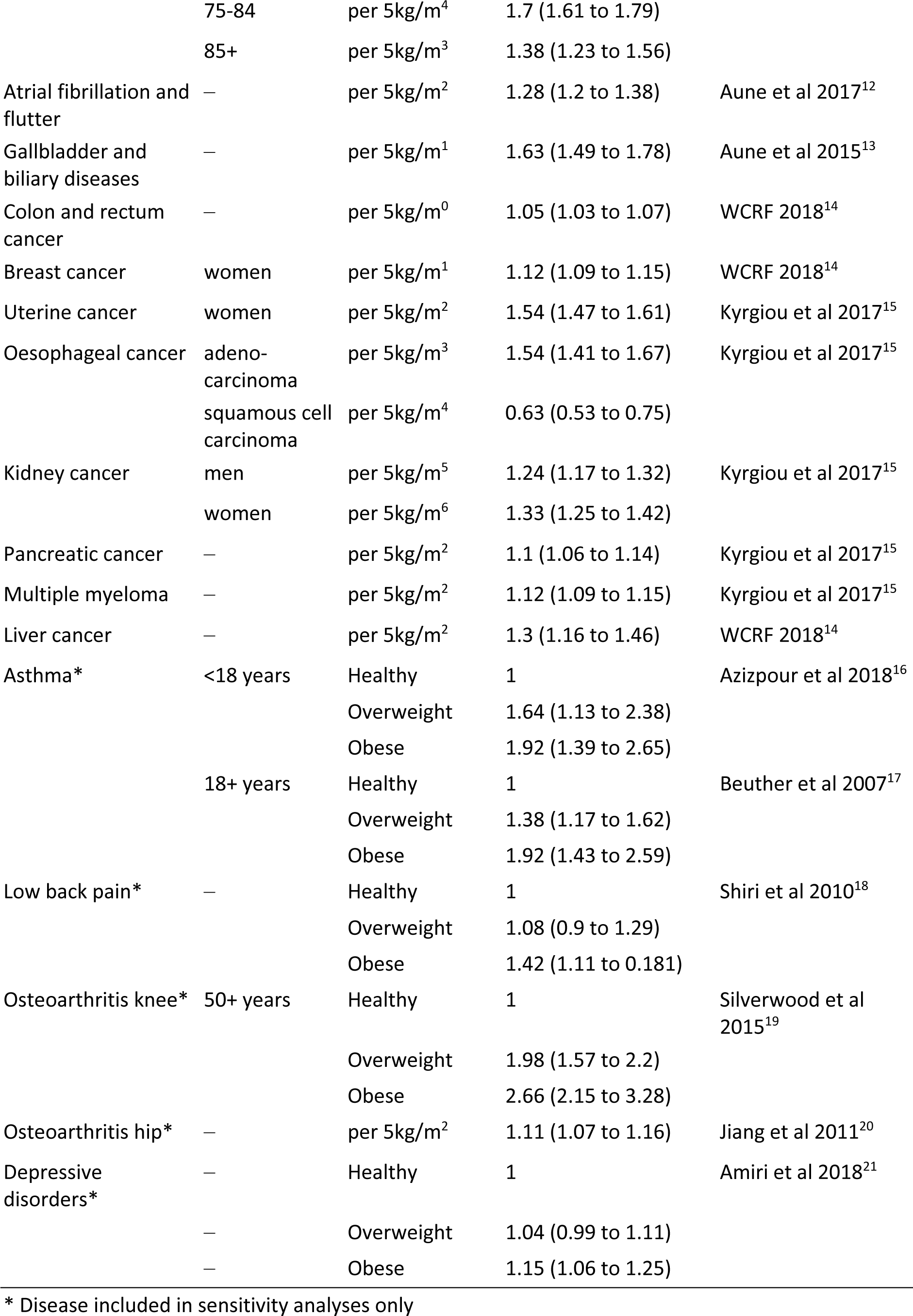
Relative risks of modelled obesity-related diseases.

**Table S5.**
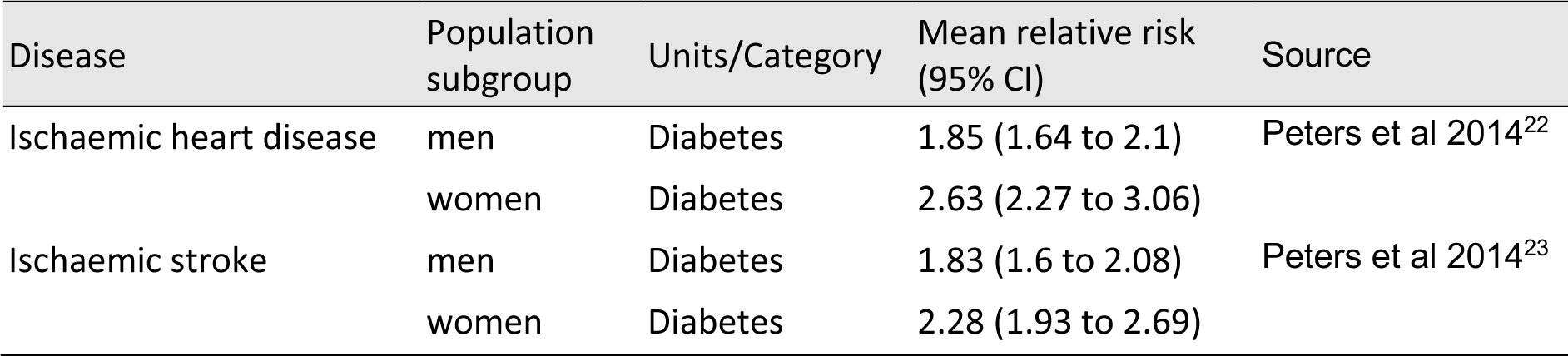
Relative risk of ischaemic heart disease and ischaemic stroke in people with type 2 diabetes.

### 10. Utility weights

**Table S6.**
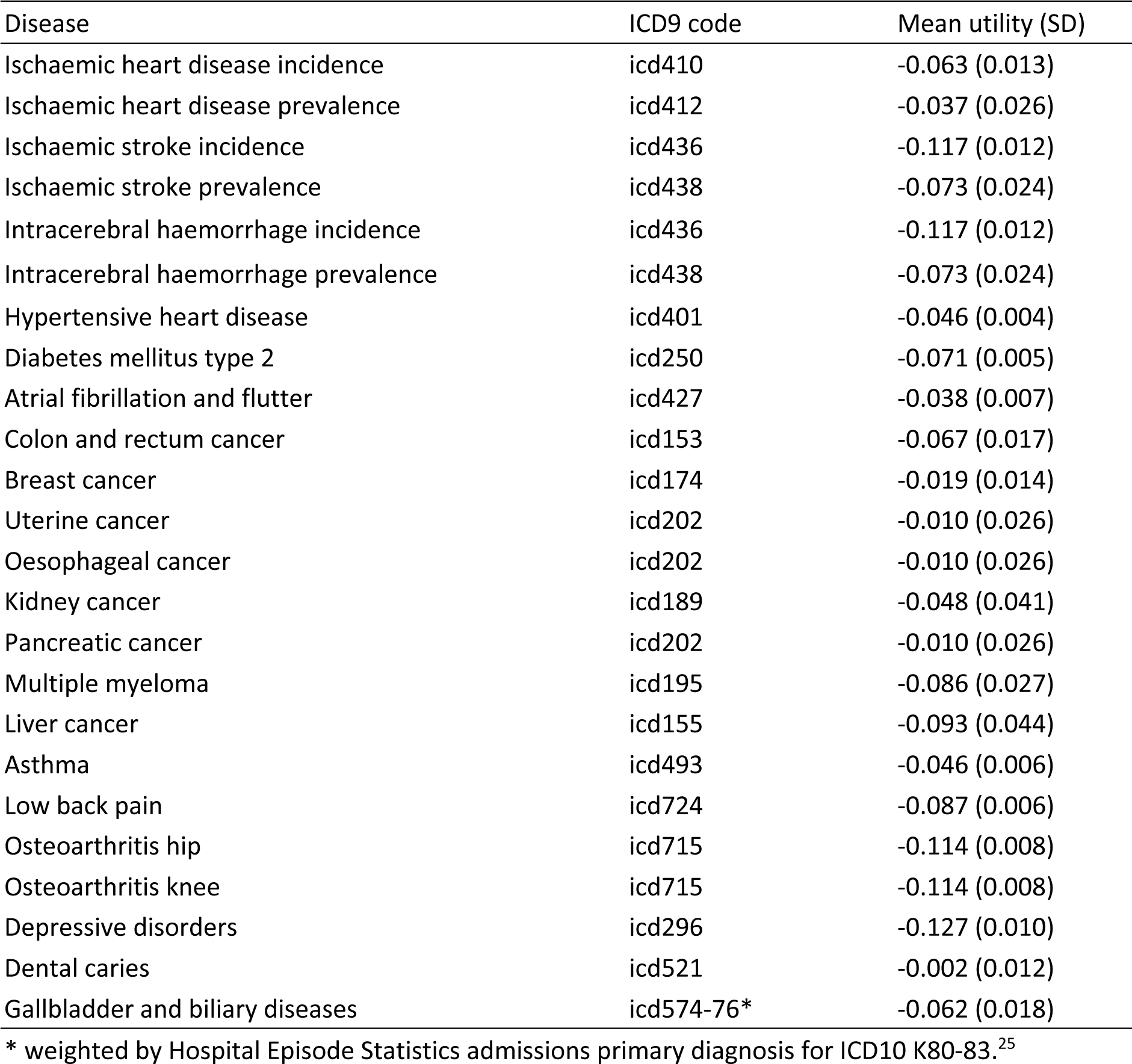
Disease-specific utility weights, estimated for the UK by Sullivan et al^24^.

**Table S7.**
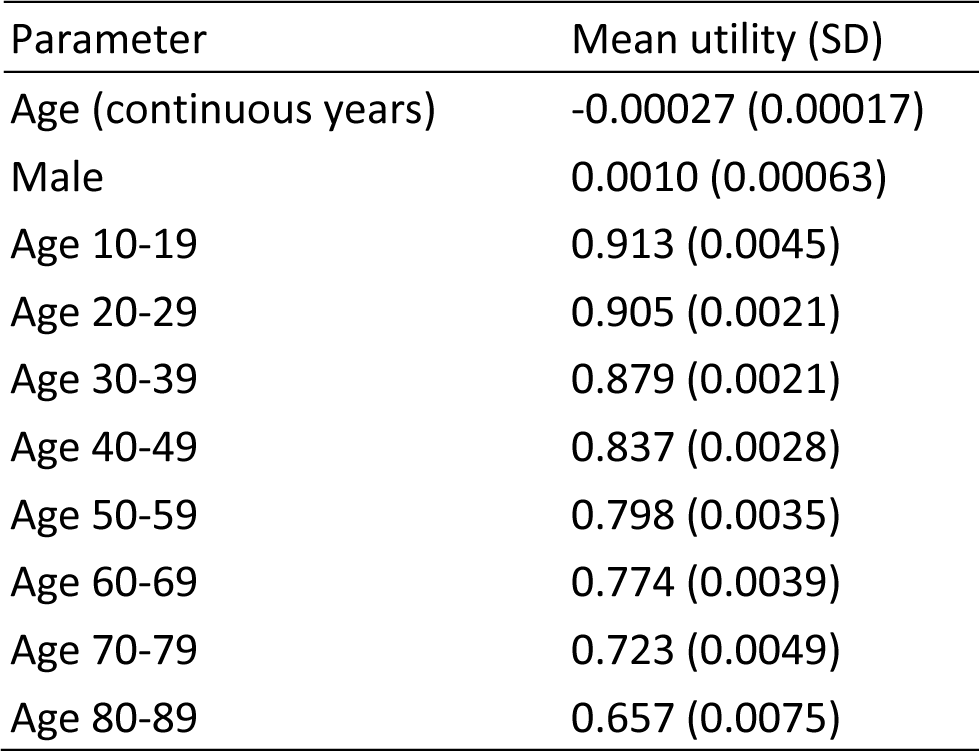
Parameters for estimating background utility weight, estimated for the UK by Sullivan et al^24^.

### 11. Disease costs

**Table S8.**
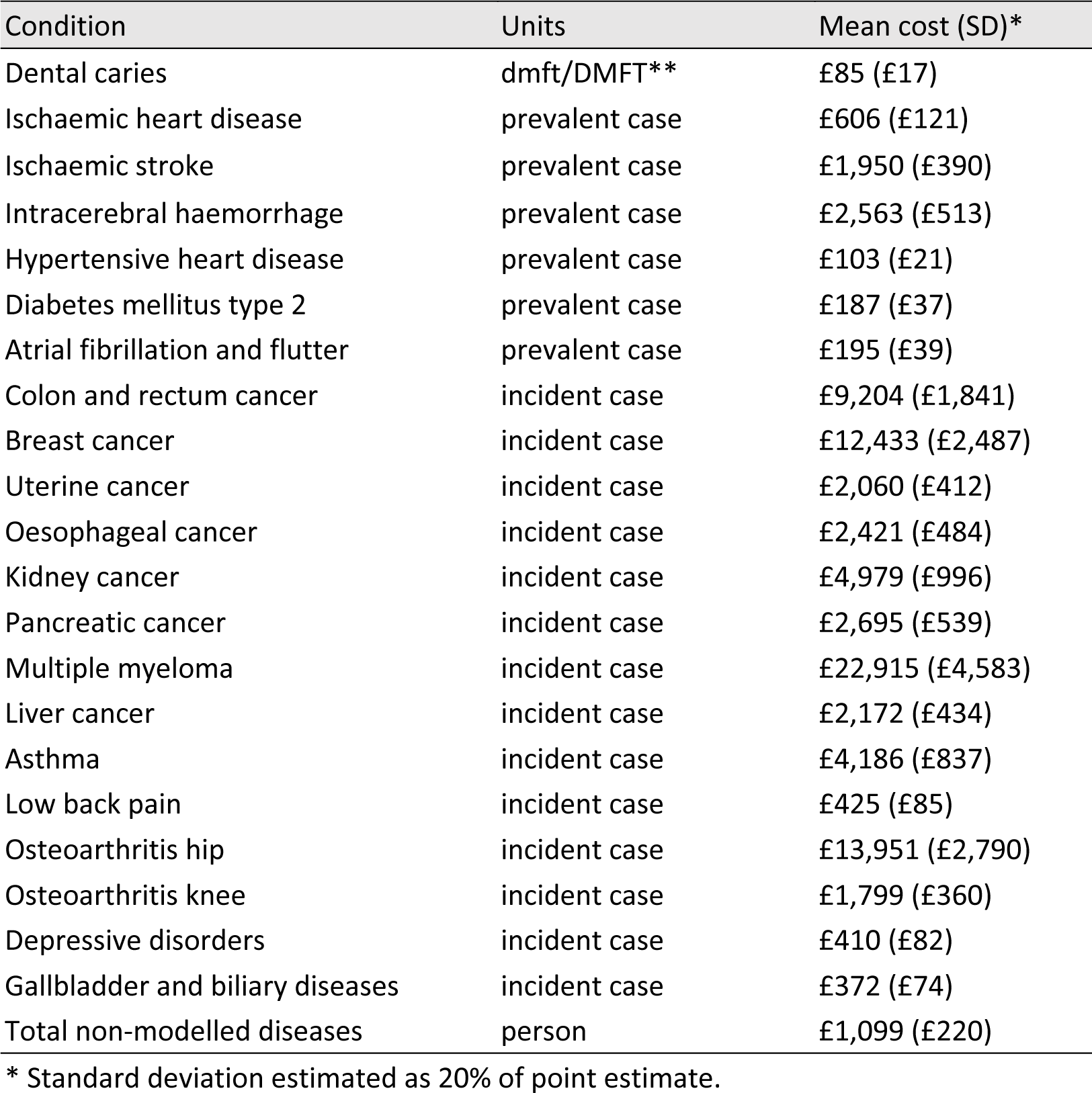
Costs of treatment in the National Health Service.

## Text S2 – Additional results

**Figure S1.**
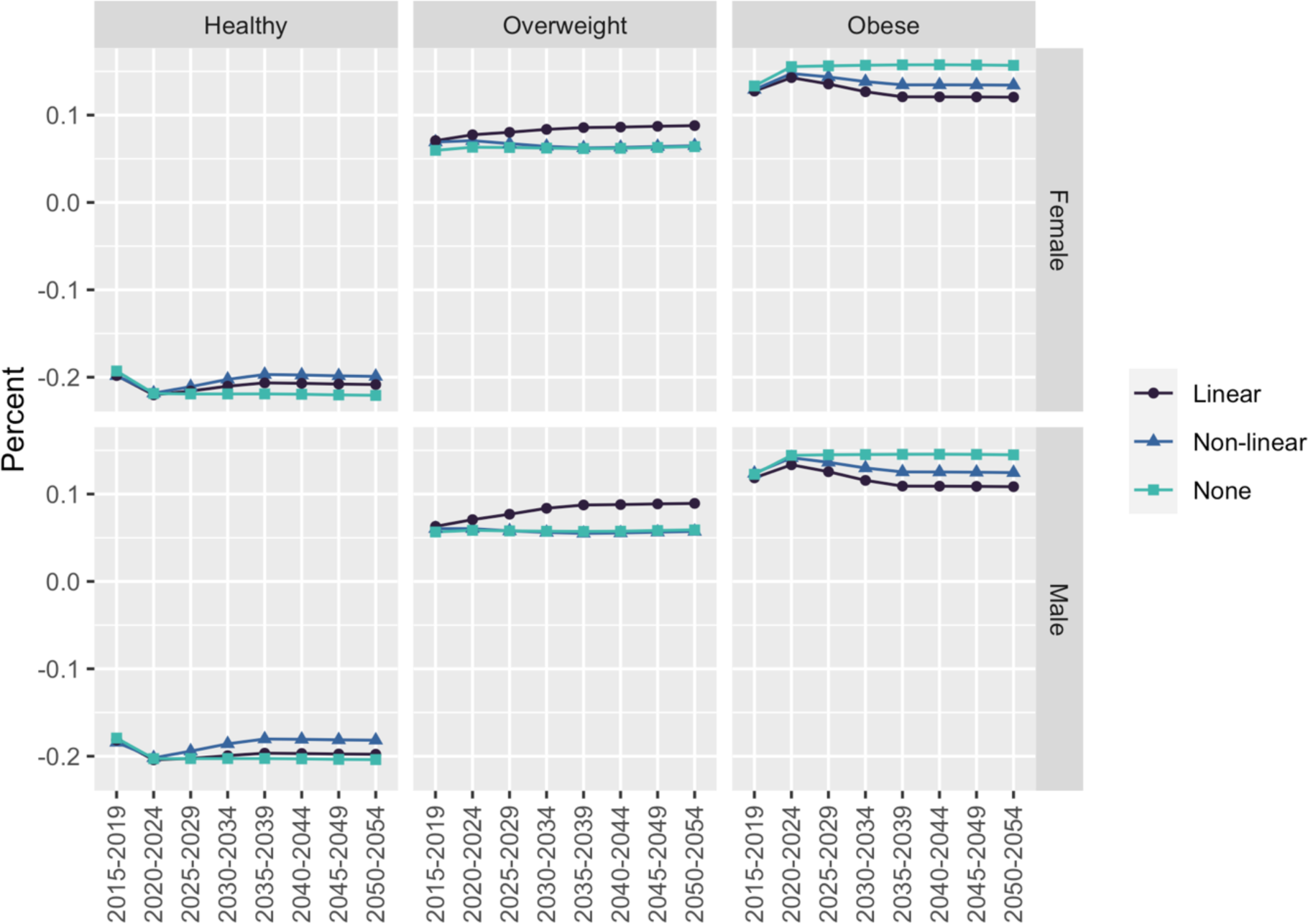
Sensitivity of predicted prevalence of overweight and obesity to the choice of BMI projection model (described in Table 1)

**Figure S2.**
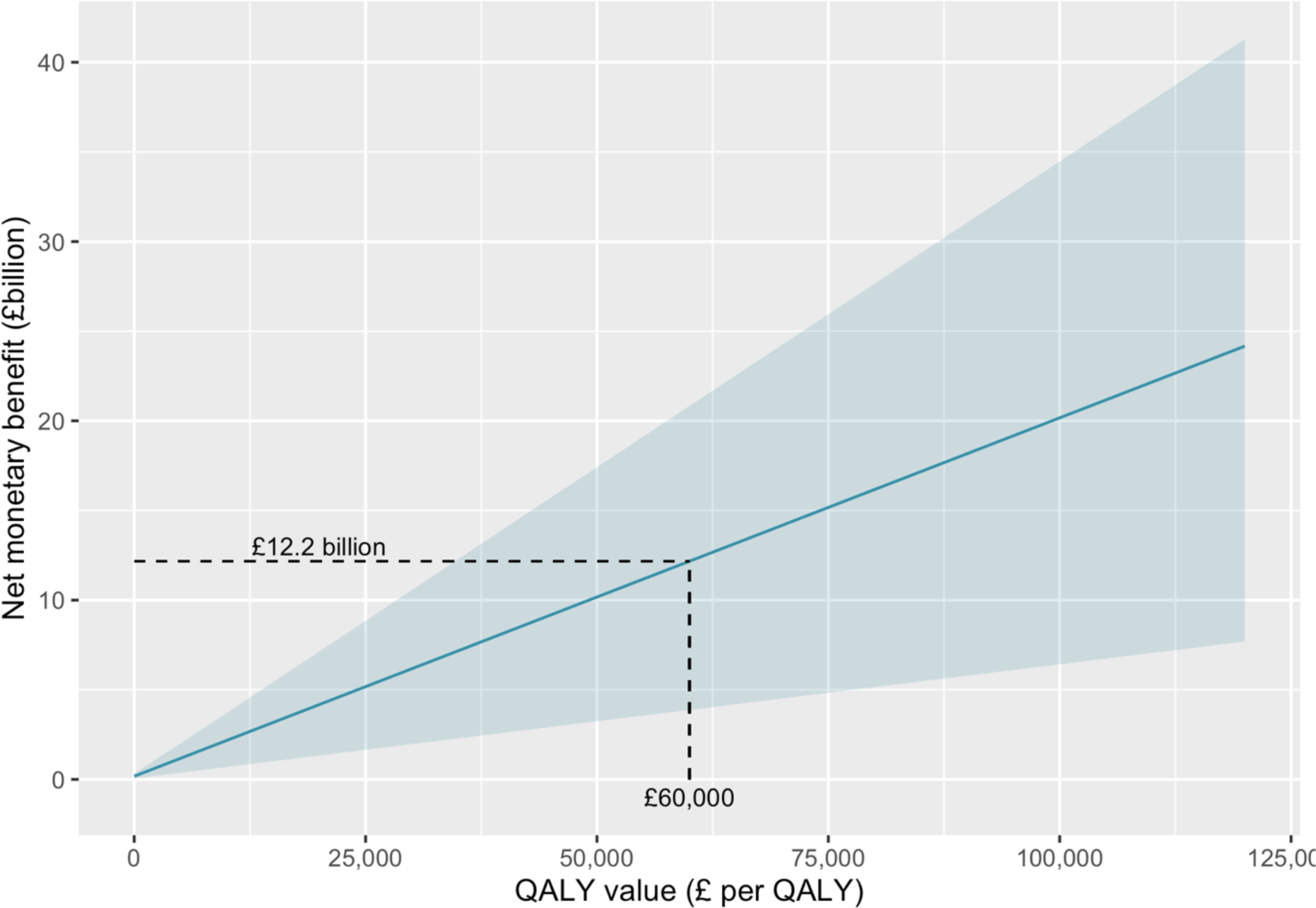
Net monetary benefit against the willingness-to-pay threshold (UK Treasury recommends a value of £60,000 per QALY)

