## Supplementary material for "Population health and health sector cost impacts of the UK Soft Drinks Industry Levy: a modelling study": Text S2

### Text S2 – Additional results

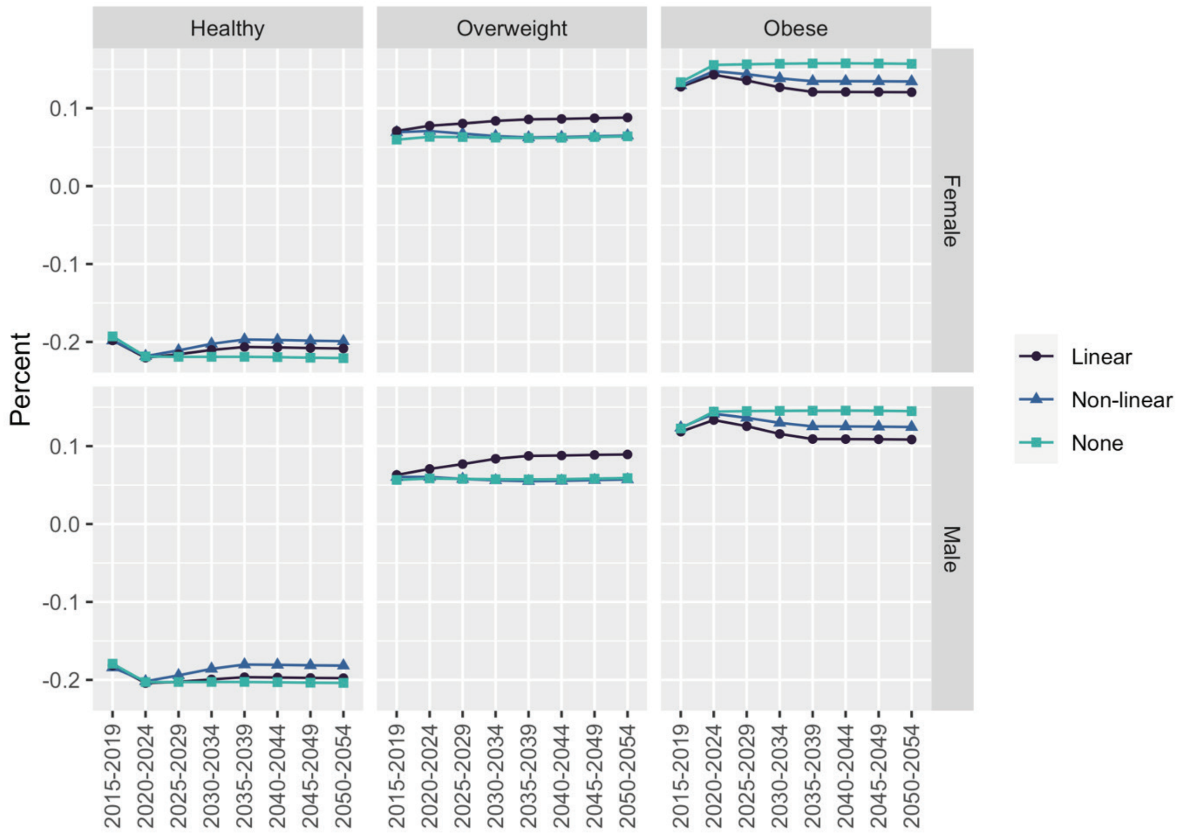


*Figure S1 Sensitivity of predicted prevalence of overweight and obesity to the choice of BMI projection model (described in Table 1)*

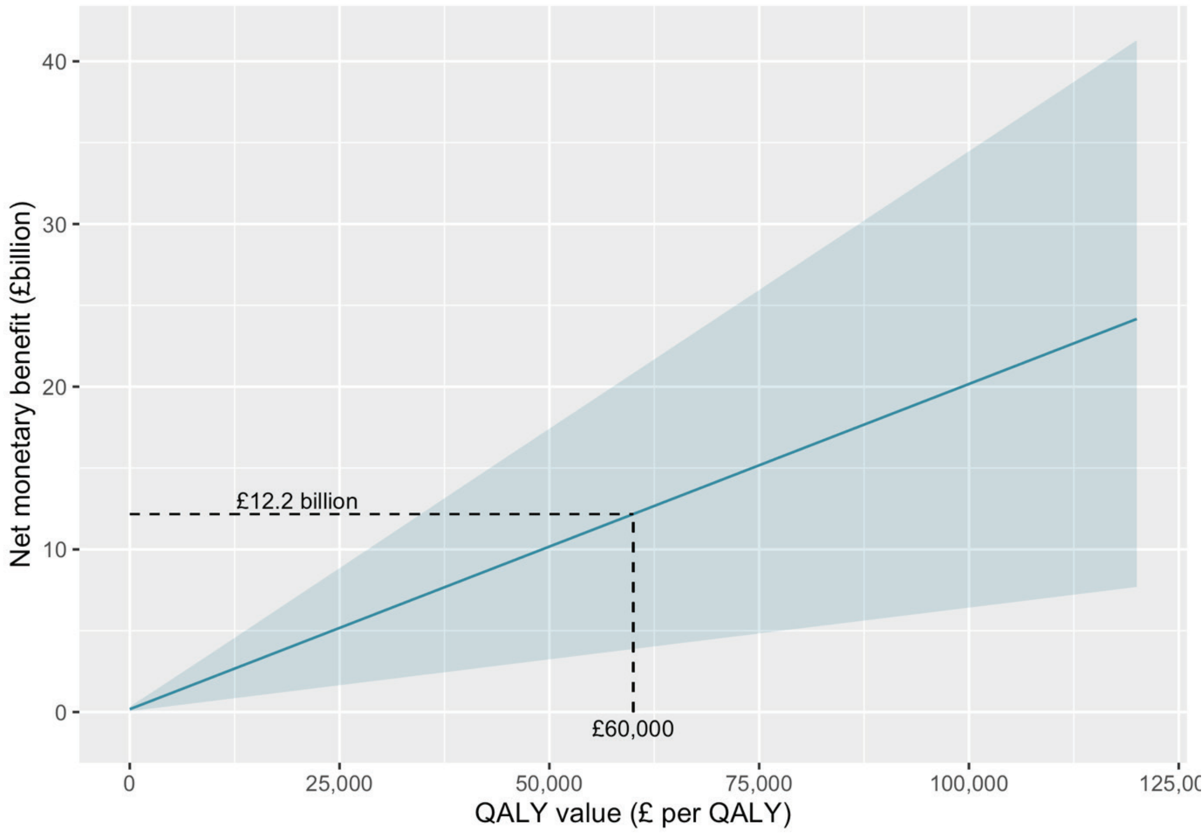


*Figure S2 Net monetary benefit against the willingness-to-pay threshold (UK Treasury recommends a value of £60,000 per QALY)*
